# Copy Number Variant Duplications Associated with Essential Tremor

**DOI:** 10.1101/2025.10.21.25337184

**Authors:** Miranda Medeiros, Calwing Liao, Allison A. Dilliott, Jay P. Ross, Veikko Vuokila, Farah Aboasali, Charles-Etienne Castonguay, Dan Spiegelman, Franziska Hopfner, Carles Vilariño-Güell, Elena García-Martín, Hortensia Alonso-Navarro, José A. G. Agúndez, Félix Javier Jiménez-Jiménez, Pau Pastor, Alex Rajput, Ali Rajput, Günther Deuschl, Gregor Kuhlenbäumer, Simon L. Girard, Sali M. K. Farhan, Patrick A. Dion, Guy A. Rouleau

## Abstract

**Background:** Essential tremor (ET) is a complex neurological disorder with a strong genetic basis, yet there remains a disparity between the estimated heritability and the currently known genetic risk of ET. This missing heritability of ET has led to a lack of appropriate treatments and has exacerbated the misdiagnosis of patients.

**Methods:** To address the missing heritability of ET, we called copy number variants (CNVs) in a large cohort of ET patients (n=1,853) and unaffected controls (n=10,336). CNVs were called from single nucleotide polymorphism (SNP) microarray data using PennCNV and QuantiSNP and only rare CNVs (frequency < 1%) intersecting protein coding regions of the genome were retained for analysis. To investigate whether CNV occurrence was associated with ET, analyses were conducted on global burden, pathogenicity burden, gene set enrichment, and gene burden.

**Results:** Global duplication burden by number of CNVs (OR = 1.71 [1.30-2.23], p = 9.8×10^-5^), CNV length (OR = 1.08 [1.02-1.15], p = 7.1×10^-3^), and number of genes affected by CNVs (OR = 1.11 [1.05-1.18], p = 5.8×10^-4^) were all significantly elevated in ET patients compared to controls. Duplications sized 100kbs-500kbs largely explained these associations (number of CNVs: OR = 2.14 [1.54-2.94], p = 3.95×10^-6^; number of genes: OR = 1.15 [1.06-1.23], p = 3.8×10^-4^). Across gene-sets, duplications affecting Mendeliome genes (OR = 2.01 [1.36-2.91], p = 3.4×10^-4^), genes highly expressed in the brain (OR = 1.80 [1.27-2.46], p = 5.06×10^-4^), and genes expressed in the cerebellum (OR = 1.16 [1.06-1.27], p = 7.31×10^-4^) were significantly enriched for CNVs in ET patients compared to controls. Finally, gene-based burden testing indicated that duplications involving *ZNF813* were significantly less frequent in ET patients than in controls (OR = 0.0411 [0.0026-0.66], p = 2.22×10^-5^). No associations with deletion events were observed.

**Conclusions:** Our results point to rare copy number duplications mapping to protein coding regions of the genome as likely contributors to ET genetic risk. However, specific susceptibility genes could not be reliably identified, highlighting the need for further studies to clarify the genetic architecture of ET.

## Introduction

Essential tremor (ET) is the most common movement disorder globally, affecting approximately 5% of the population by the age of 65 years.^1^ ET is a complex and progressive neurological disorder, and its risk continues to increase with age.^2^ Patients with ET suffer from tremors during voluntary movement occurring primarily in the hands, head, and voice, resulting in a significantly diminished quality of life.^1,3^ Affected individuals often rely on caregivers for physical and emotional support, experience depression and anxiety, and struggle with daily living activities.^3^

With a quarter of an ET patient’s first-degree relatives also likely affected,^4^ the strong genetic component of the condition is evident. However, no clinically actionable genes for ET are currently known.^2^ Unfortunately, the lack of specific genetic markers and understanding of ET pathophysiology means that many patients with tremor symptoms are misdiagnosed, with up to 50% of cases being mistaken for disorders like Parkinson’s disease.^5^ This prevents ET patients from obtaining the proper treatments they require, exacerbating the burden to the healthcare system, and highlighting the need to establish the genetic architecture of ET.^3^ Given its strong genetic basis, with an estimated heritability ranging from 45%-90%,^4^ many studies have attempted to identify the genes responsible for ET.^6^ However, proposed ET variants tend to be limited to single families and typically do not replicate across studies.^6^ Many strategies, such as twin studies, linkage studies, genome association studies (GWAS), as well as exome and genome sequencing approaches have been utilized to identify ET genetic risk factors. However, while GWAS have identified some associations, they do not fully explain the phenotype, and individual variants identified alone tend to carry very little explanatory power contributing modestly to the overall genetic risk. At the same time, rare variant studies have primarily shown that rare variants in ET are often private to specific pedigrees, lacking generalizability.^6^ Ultimately, the current gap between known ET genetic risk and estimated heritability is wide and has prompted us to interrogate other types of variants that have been largely unexplored in ET to address the missing heritability of the disorder.

Copy number variants (CNVs) are large structural variants, including deletions or duplications of genetic material, ranging from hundreds to millions of base pairs in size. These variants can encompass parts of or entire genes, making them likely to be damaging.^7^ It is well known that CNVs can contribute major risk to neurodevelopmental disorders such as autism spectrum disorder (ASD).^8^ However, CNVs have also been implicated as risk factors in movement disorders like Parkinson’s disease.^9,10^ In some exceptional cases, repeat expansions have even been reported to reside within CNVs, adding to the potential complexity of structural events and their consequences on disease risk.^11^ Though rare CNVs have been associated with rare disorders previously, they have also been shown to play a role in common disorders, chiefly by increasing disease risk and causing earlier ages of onset.^12^ Moreover, in the movement disorder space, CNVs have been able to increase diagnostic yield for dystonia patients,^13^ and in terms of later onset-disorders, such as in Alzheimer’s disease, associations between disease status and CNVs have been reported.^14^

However, largely due to the innate challenges in accurately detecting them, CNVs remain an understudied source of genetic risk compared to other variant types, namely single nucleotide variants (SNVs).^15^ To date, large-scale CNV studies have yet to be performed in ET. As such, we predicted that a missing source of ET genetic risk could be attributed to CNVs.

In this study, we leveraged a large cohort of ET patients (n = 1,853) and unaffected controls (n = 10,336) to explore whether rare CNVs mapping to protein-coding regions contribute to the genetic landscape of the condition using a series of robust statistical tests.

## Methods

### CNV calling

#### Cohort

A cohort of ET samples (n=1,853; 54% female; 46% male; average sample age [standard deviation (SD)]: 65.8 [15.5] years; average age of onset [SD]: 41.4 [21.9] years) were genotyped using the Infinium Global Screening Array (Illumina, Inc.). ET patient cohort and diagnostic criteria were outlined previously.^16^ Briefly, diagnoses were made based on established criteria and reviewed by movement disorder specialists or senior neurologists to ensure accuracy. Patients provided informed consent, and ethical approval was obtained from relevant ethics committees. Exclusion criteria included a history of other neurological disorders, such as Parkinson’s disease, or conditions that could mimic ET, such as exaggerated physiological tremor, dystonia, or psychogenic tremors. Patient samples were sourced from Canada (n = 613) and Europe (n = 1,240). A neurologically healthy cohort from the Québec population database CARTàGENE (n=10,336; 52% female; 48% male; ages: 40-69 years old) were genotyped using the Infinium Global Screening Array (Illumina, Inc.) and used as controls.^17^ Further details about CARTàGENE are described in *Awadalla et al*.^17^

#### Genotyping and Sample Quality Control

Raw genotyping intensity files were loaded onto GenomeStudio v.2.0 (Illumina, Inc.) and quality controlled at the individual level and variant level, as previously described.^18^ Only single nucleotide polymorphisms (SNPs) common to both case and control datasets were used in analyses and CNV calling. Sample ancestry was estimated using principal component analysis (PCA) using PLINK v.1.9, with the 1000 Genomes Project as a reference panel.^19,20^ As 94% of samples clustered with European samples of the 1000 Genomes Project, only individuals with inferred European ancestry were included. The Kinship-based INference for Gwas (KING) algorithm was used to retain unrelated individuals (pairwise second-degree relatedness threshold).^21^ Further, samples with an abnormally high number of CNV calls (>150 CNVs per sample) were removed as previously described.^22^ Standard thresholds of Log R ratio (LRR) standard deviation, B Allele Frequency (BAF) drift, and Waviness Factor (WF) were applied (3×SD) to remove poor quality samples.^23^ After sample level quality control, 1,204 ET cases and 9,549 controls remained. Quality control steps are outlined in Figure 1. Based on our sample size and the focus on rare events (disease allele frequency of ≤ 1%), the study had 80% power to detect an odds ratio of 1.76 or greater as calculated by the University of Michigan Genetic Association Study Power Calculator. Smaller or more modest associations may therefore have gone undetected.

**Figure 1.**
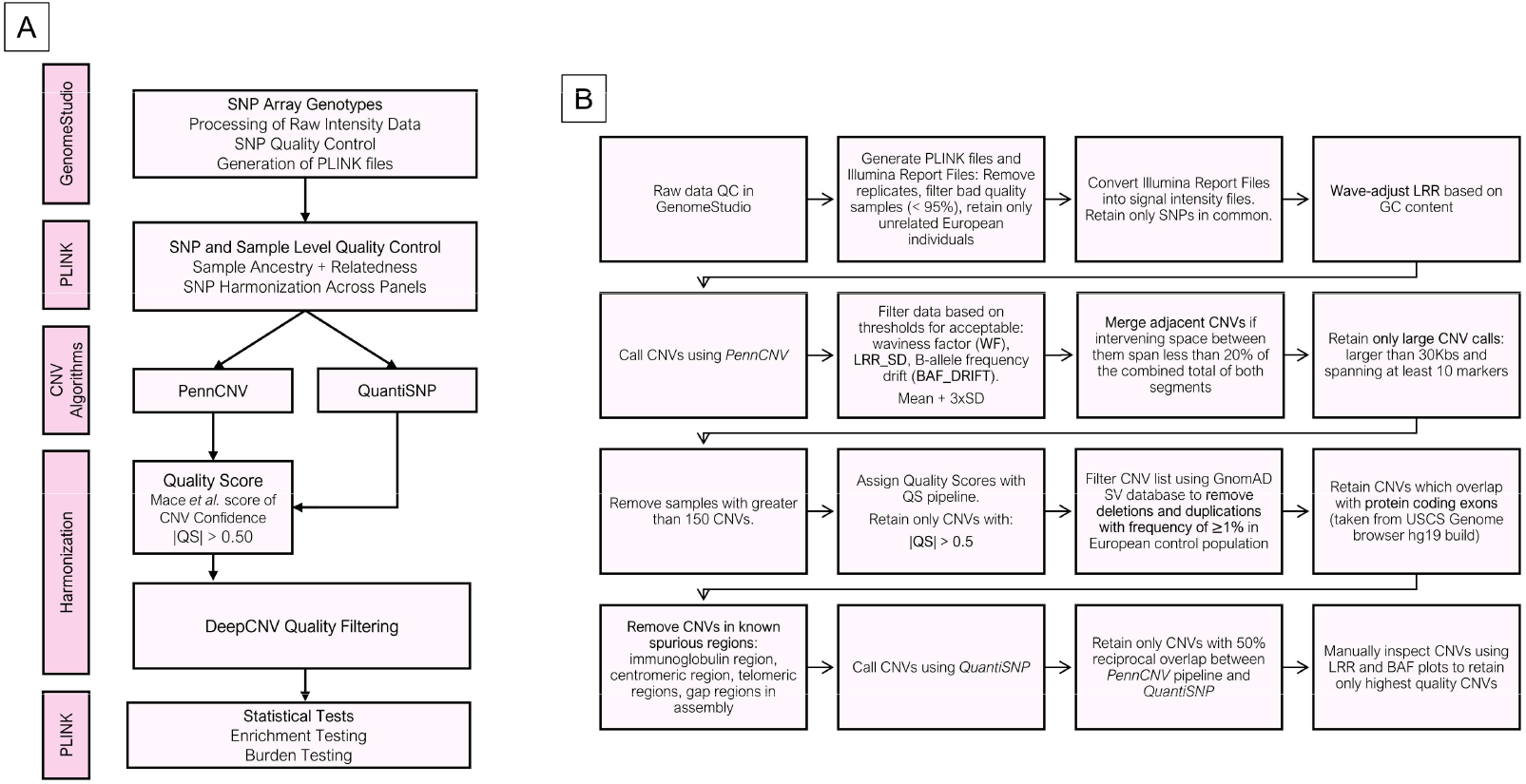
Copy number variant processing workflow. (A) The copy number variant (CNV) calling pipeline applied to Infinium Global Screening Array genotyping data from individuals with ET (n=1,853) and controls (n=10,336). (B) Quality control steps applied to the called CNVs.

#### CNV level quality control

CNVs were called using PennCNV^24-26^ and QuantiSNP^27^ (Figure 1). For quality control at the CNV level, CNV calls were removed if they were less than 30kb in length or spanned fewer than 10 SNP markers.^23^ To mitigate false positive CNV calls detected using PennCNV, the Quality Score (QS) model outlined by Macé et al.^28^ was applied to identify high quality CNV calls. The QS captures the agreement between calls generated from PennCNV compared to two other CNV calling algorithms: CNVpartition^29^ and QuantiSNP^27^. High quality consensus calls were generally considered as CNVs with |QS| > 0.50.^28^ Rare CNVs were defined by the GnomAD Structural Variant dataset v4.1 (European AF < 1%) for deletion and duplication events that reciprocally overlapped CNVs by at least 50%.^30^ Only rare CNVs were retained. Any CNV call overlapping with poorly mapped regions (ENCODE Blacklist) of the genome (UCSC GRCh37/hg19) including centromeric and telomeric regions were removed.^31^ To validate our approach, CNVs were also called by QuantiSNP.^27^ Only CNVs called from PennCNV that reciprocally overlapped with QuantiSNP CNVs by at least 50% were retained. Next, only high-confidence CNVs identified through DeepCNV were retained.^32^ The DeepCNV tool classifies CNV quality through BAF and LRR plots, ultimately determining CNV confidence based on consistent LRR deviation from 0 and specific BAF clustering patterns. Finally, only rare CNVs that intersected at least one protein coding exon were retained.^33^ The CNV calling pipeline described is shown in Figure 1.

### Statistical analyses

#### Global burden analyses

Analyses began with global burden tests to assess if there was: a greater number of CNVs, a greater CNV length, and a greater number of genes mapped within CNVs in ET cases compared to controls across the genome. Global CNV burden was assessed through logistic regressions using different burden metrics: CNV count, CNV length (measured by 100kbs units), CNV gene count. Sex, the first 10 principal components (PCs), and LRR standard deviation (LRR_SD; used as a quality metric) were included as covariates (ET_Status ∼ Burden_metric + sex + PC1-10 + LRR_SD). Separate regressions were performed for all deletions and duplications (Bonferroni threshold p-value (α/n): 0.05/3). Independent regressions for each size group were also conducted separately for deletions and duplications segregated by size (CNVs: >100 kbs, 100–500 kbs, 500 kbs–1 MB, and >1 MB; Bonferroni threshold p-value (α/n): 0.05/8).

#### Global gene-wise burden analyses

To assess the burden of CNVs at the gene level globally, we performed gene-wise burden analyses, comparing the number of CNVs affecting each gene individually in ET cases and comparing it to controls using Fisher’s exact tests. Each gene with at least one exon encompassed by a CNV was analyzed independently via a Fisher’s exact test to assess the burden of CNVs. A Bonferroni significance threshold (α/n) of 0.05/1891 was applied to deletions and duplications. Odds Ratios (ORs) for Fisher’s exact tests with 0 counts were manually calculated according to Altman (1991).^34^ This approach was applied to all genes globally, as well as to the subset of genes belonging to any significantly enriched gene-set result, to attempt to identify the genes driving the gene-set signal.

#### Gene-set enrichment analyses

Enrichment analyses were conducted using a logistic regression model to identify associations between ET status and the number of CNVs affecting genes belonging to a given gene-set, adjusting for sex, PCs 1-10, and LRR standard deviation (ET_Status ∼ Gene_Set + sex + PC1-10 + LRR_SD). We determined the number of CNVs encompassing genes belonging to each given gene-set for deletions and duplications separately in the ET cohort and the control cohort. Gene-sets included: genes previously associated to ET, constrained gene identity score of mammalian orthologs (GISMO)^35^ genes, constrained GISMO-missense genes, constrained sHet^36^ genes, Gnomad v2.1.1 missense constrained genes, Gnomad v2.1.1 loss of function constrained genes,

Mendeliome genes, zinc finger protein genes (since several CNVs affecting zinc finger genes were called across cases and controls), transcription factor genes, genes with elevated or exclusive expression in the brain, genes which are expressed in the cerebellum (a key region for ET), synaptic genes, ClinGen triplosensitive genes, ClinGen haploinsufficient genes, rare CNV (rCNV) triplosensitive genes, rCNV haploinsufficient genes, and neurodegenerative genes. Exact details about how these gene-sets were built are described in eMethods. Regressions were performed for deletions and duplications separately (Bonferroni threshold p-value (α/n): 0.05/30).

For any significant gene-set signals, subsequent gene-level burden analyses were performed using Fisher’s exact tests. Tests compared the number of CNV events between cases and controls affecting genes within significant gene-sets as described in the gene-wise burden section of the Methods. Bonferroni thresholds for these tests were gene-set specific (α/n): 0.05/(number of genes in gene-set).

#### CNV burden in genomic disorder regions analyses

Burden analyses were done to understand the distribution of CNVs in and outside of known CNV-disease association hotspots (genomic disorder regions) between ET patients and controls. We compiled a reference list of genomic disorder regions^37^ and assessed CNV overlap with these regions using a ≥50% reciprocal overlap threshold. For deletions and duplications separately, we constructed 2×2 contingency tables comparing the number of CNVs overlapping genomic disorder regions versus CNVs outside genomic disorder regions across cases and controls. Fisher’s exact tests were then used to determine whether the distribution of CNVs within genomic disorder regions compared to outside of genomic disorder regions differed significantly between ET cases and controls.

#### Pathogenic prediction scoring burden analyses

Using X-CNV, rare CNVs affecting at least one exon were annotated with scores from the most predictive pathogenic metrics of coding and genome-wide features: logistic regression (LR) score, Variant Effect Scoring Tool (VEST3) score, Functional Analysis Through Hidden Markov Models (FATHMM) score, probability of being loss-of-function intolerant (pLI) score, Combined Annotation Dependent Depletion (CADD) score, and the X-CNV combined meta-voting prediction (MVP) score.^38^ Each CNV was annotated with each pathogenic score metric separately. To better understand the characteristics of the CNVs belonging to ET patients and controls, we calculated the mean score across deletions and duplications for each of the six-pathogenicity metrics (LR score, VEST3 score, FATHMM score, pLI, CADD score, and MVP score) for each individual. This yielded six separate average pathogenicity scores per sample (one for each metric) which were then independently tested for association with case-control status via logistic regressions. The model used served to identify associations between ET status and a given averaged pathogenicity score, adjusting for sex, PCs 1-10, and LRR standard deviation (ET_Status ∼ Average_Pathogenicity_Metric + sex + PC1-10 + LRR_SD). Regressions were performed for deletions and duplications separately (Bonferroni threshold p-value (α/n): 0.05/12).

## Results

### CNV calling

Following sample and CNV level quality control, a total of 1,204 ET patients and 9,549 controls were included in our analyses. Of these, 244 ET patients (22.42%) were found to carry 143 deletions and 180 duplications, while 2,402 unaffected controls (25.15%) were found to carry 1,348 deletions and 1,709 duplications using PennCNV^24-26^ and QuantiSNP^27^ (Supplementary Figure 1). All deletions had a copy number of 1, and all duplications had a copy number of 3.

### Global burden

To investigate whether there was a global increase in the number of CNVs, CNV length, and the number of genes mapped within CNVs in ET patients compared to controls, we conducted global burden analyses using logistic regressions across all rare CNVs affecting protein coding regions. The analyses revealed no significant associations (p-value >0.05) between ET status and any burden metric across deletions. However, for duplications, the total number of duplications (OR = 1.71 [1.30-2.23], p = 9.8×10-5), total length of duplications in units of 100kbs (OR = 1.08 [1.02-1.15], p = 7.1×10-3), and number of unique genes affected by duplications (OR = 1.11 [1.05-1.18], p = 5.8×10-4) were significantly associated with ET status (Figure 2).

**Figure 2.**
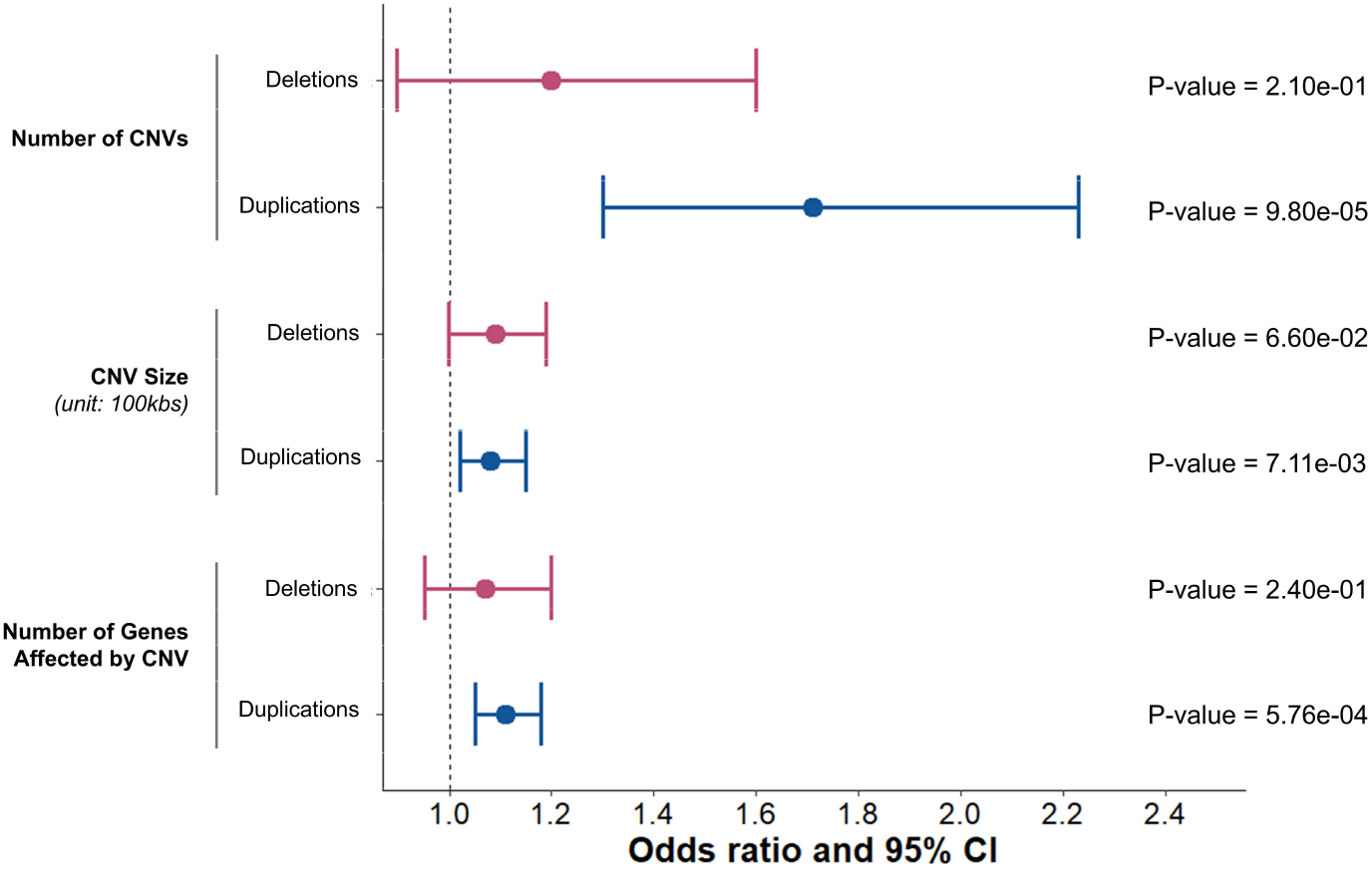
Global burden analyses of rare copy number variants encompassing protein coding regions in individuals with ET (n=1,204) compared to controls (n=9,549). Forest plot depicting logistic regression results from tests of ET status with number of genes affected by copy number variants (CNVs), number of CNVs, and CNV size (measured in units of 100kbs) between ET patients and controls are shown. Odds ratios and 95% confidence intervals for each burden metric regression analyses are shown in red for deletions and in blue for duplications. P-values are also displayed. Bonferroni adjusted p-value cut-off: 0.05/3.

We next categorized CNVs into four size-based bins (<100 kbs, 100 kbs–500 kbs, 500 kbs–1 MB, and >1 MB) to investigate potential global burden signals, focusing on both the total number of CNVs and the number of genes affected by these CNVs. No significant associations were found between ET status and any deletion size subset. The only significant associations of a size subset with ET status were seen in duplications in the size range of 100kbs-500kbs for total number of CNVs (OR = 2.14 [1.54-2.94], p = 3.95×10^-6^), and total number of genes (OR = 1.15 [1.06-1.23], p = 3.8×10^-4^) (Figure 3).

**Figure 3.**
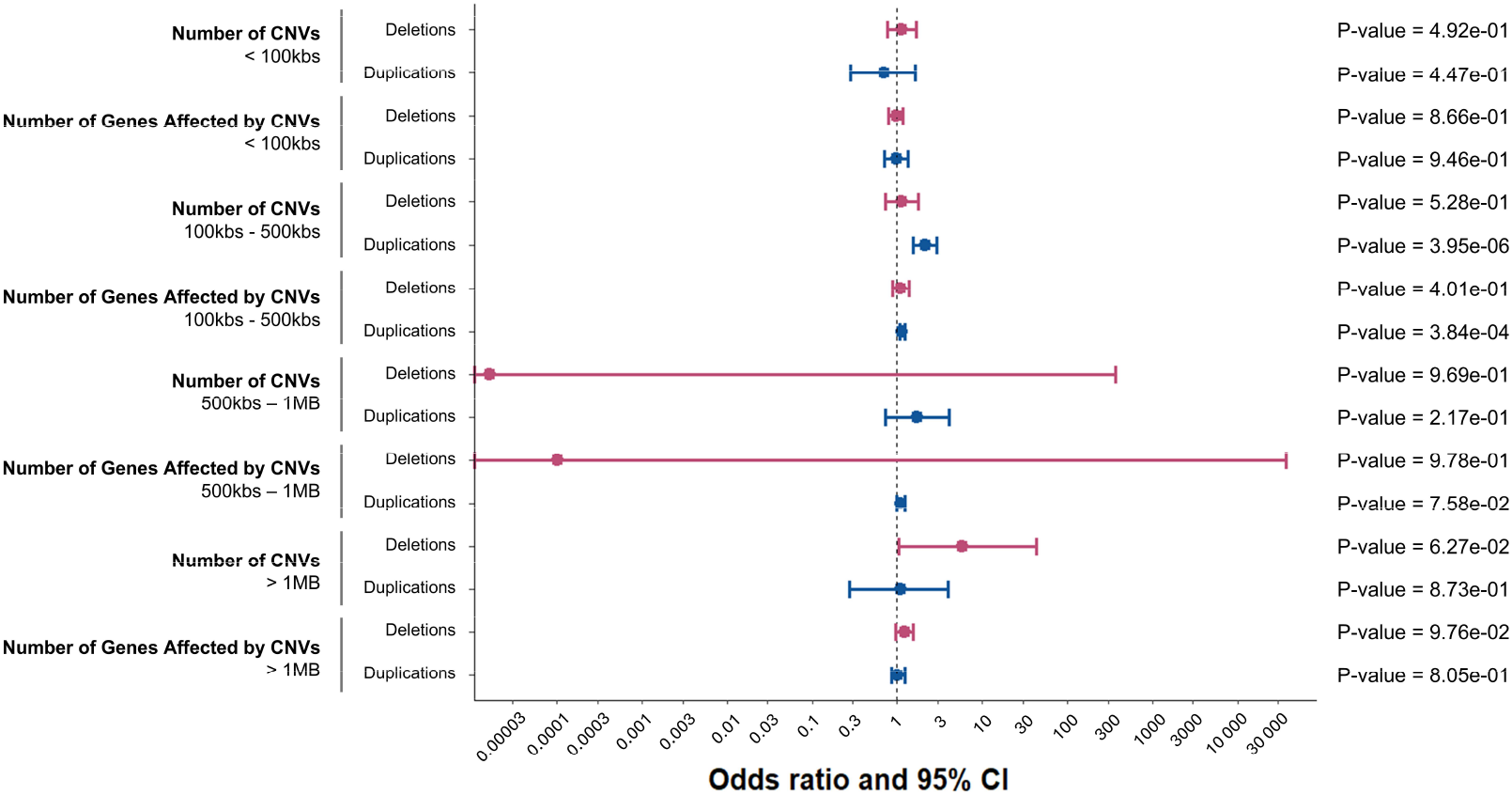
Global burden analyses of rare copy number variants encompassing protein coding regions segregated by size. Forest plot depicting logistic regression results from tests of ET status with number of genes affected by copy number variants (CNVs) and number of CNVs between ET patients and controls are shown for different CNV size cut-offs. CNVs are divided in 4 subsets based on size, these are CNVs: < 100kbs, 100kbs – 500kbs, 500kbs – 1MB, and > 1MBs. Odds ratios and 95% confidence intervals for each burden metric regression analyses are shown in red for deletions and in blue for duplications. Due to low sample size, the models for duplications < 100 kbs did not fit and as such are not displayed. P-values are also displayed. Bonferroni adjusted p-value cut-off for deletions: 0.05/8. Bonferroni adjusted p-value cut-off for duplications: 0.05/6.

### Global gene-wise burden

To identify potential ET risk genes, we focused our analysis on the individual genes affected by rare CNVs. CNV burden for each gene affected by a CNV was assessed independently per gene. These analyses were done using Fisher’s exact tests where the total number of CNVs encompassing a given gene was compared between cases and controls for deletions and duplications separately (eTable 1 & eTable 2). Across all genes, there were significantly fewer ET patients with duplications encompassing *ZNF813* than controls (OR = 0.0411 [0.0026-0.66], p = 2.22×10^-5^). Specifically, no ET case carried a duplication affecting *ZNF813*, whereas 95 controls did. *ZNF813* is predicted to enable DNA-binding transcription factor activity but has low tissue specificity and has relatively low expression in the brain.^39^ No other association passed multiple hypothesis correction.

### Gene-set enrichment

We next focused our analysis on a series of curated gene-sets to reduce potential noise generated by CNVs capturing genes less likely to be relevant to ET. The tested gene-sets are listed in the Methods and described in detail in the eMethods. No rare CNVs identified in individuals with ET or controls encompassed genes included in the rCNV triplosensitive, rCNV haploinsufficent, ClinGen triplosensitive, ClinGen haploinsufficent, nor neurodegenerative gene-sets. No gene-sets showed significant enrichment for deletions. Mendeliome genes (OR = 2.01 [1.36-2.91], p = 3.4×10^-4^), genes highly expressed or exclusively expressed in the brain (OR = 1.80 [1.27-2.46], p = 5.06×10^-4^), and genes expressed in the cerebellum (OR = 1.16 [1.06-1.27], p = 7.31×10^-4^) were found to be significantly enriched for duplications in ET patients compared to controls (Figure 4).

**Figure 4.**
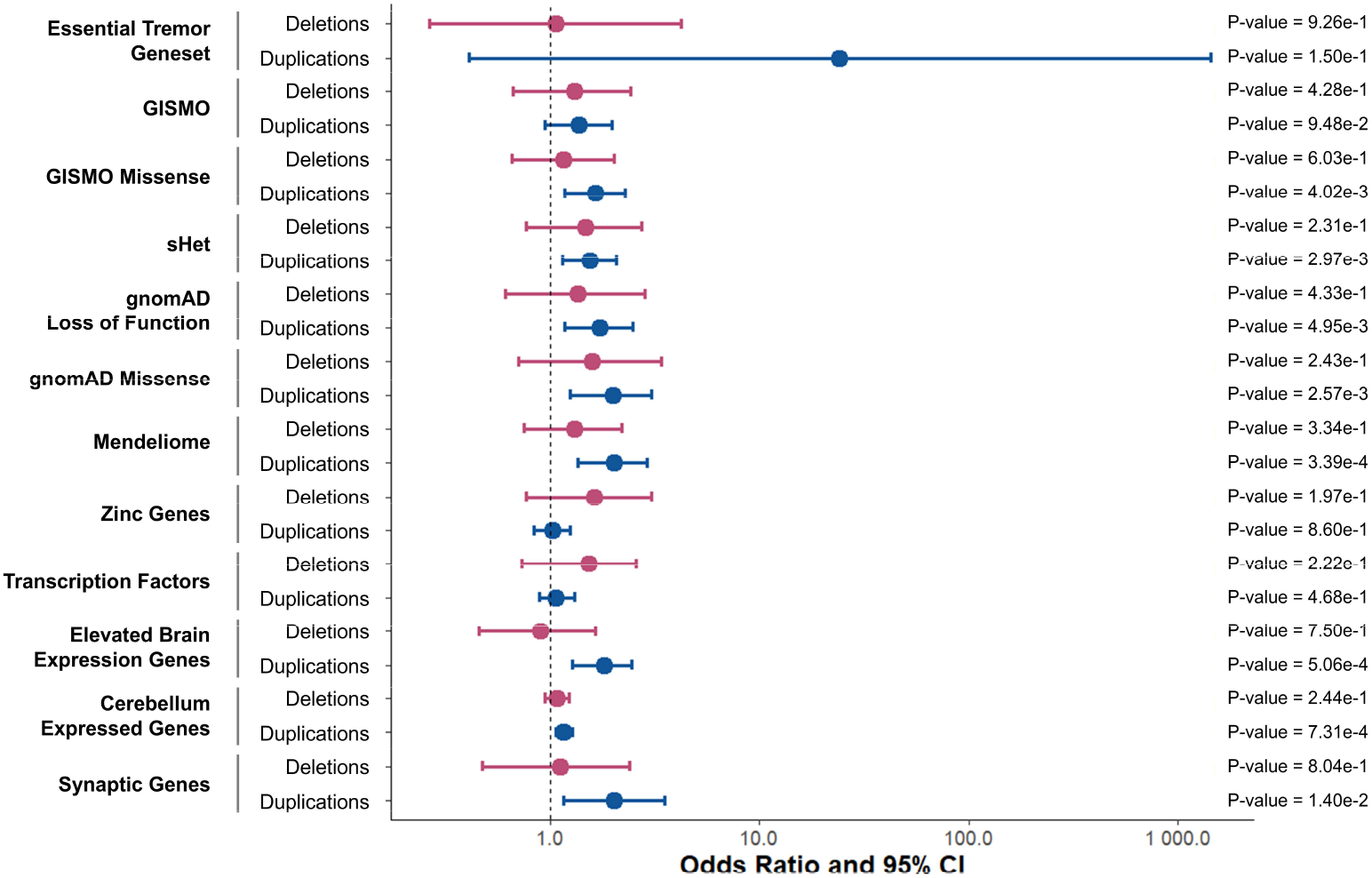
Gene-set enrichment analyses of rare copy number variants encompassing protein coding regions. Forest plot depicting logistic regression results from tests of ET status with copy number variant (CNV) counts across gene-sets are shown. Odds ratios and 95% confidence intervals for each burden metric regression analyses are shown in red for deletions and in blue for duplications. No CNVs belonging to cases nor controls possessed duplications that affected any gene belonging to the rare CNV (rCNV) triplosensitive or haploinsufficient and ClinGen triplosensitive or haploinssuficent gene-sets, nor did any CNV affect any gene in the neurodegenerative gene-set. These gene-sets are therefore not depicted in the figure. Bonferroni adjusted p-value cut-off was: 0.05/30.

We further interrogated the genes making up the significant gene-sets. For genes highly expressed and exclusively expressed in the brain, 231 genes were affected by duplications, however subsequent gene-wise burden testing revealed that no single gene carried a greater burden of duplications between cases and controls. For genes with expression in the cerebellum, 1,484 genes were affected by duplications, one of which being the *ZNF813* gene. For Mendeliome genes, we discovered that 188 genes belonging to the gene-set had been affected by duplications. However, at the gene level no single gene displayed a significant burden of duplications across Mendeliome genes.

### Genomic disorder region burden

To investigate whether CNVs contribute to ET through known regions of genomic instability and disease relevance, we tested for enrichment of rare, protein-coding CNVs within established genomic disorder regions in ET cases compared to controls. These genomic disorder regions are known hotspots for recurrent CNVs and disease associations, making the distribution of CNVs in these regions of particular interest in ET. Fisher’s exact tests were used to compare the distribution of CNVs inside versus outside these genomic disorder regions in ET cases compared to controls. No association was found for deletions (OR = 2.71 [0.651-8.502], p = 8.51e-2); however, a greater burden of duplications in disorder regions was revealed in ET cases compared to controls (OR = 2.892 [1.261-6.053], p = 6.41e-3). Specifically, 3.57% of deletions across cases corresponded to genomic disorder regions whereas 1.35% of control deletions corresponded to these regions. For duplications, 6.45% of duplications in cases were in genomic disorder regions in contrast to 2.32% of control duplications.

### Pathogenic prediction scoring burden of rare CNVs

To assess whether CNVs carried by individuals with ET were more likely to be pathogenic than controls, we compared average pathogenicity prediction scores across CNVs belonging to CNV carriers. Importantly, these analyses focused on predicted deleteriousness among individuals carrying CNVs, rather than comparing CNV presence between cases and controls. We compared averaged X-CNV^38^ logistic regression (LR) scores, Variant Effect Scoring Tool (VEST3) scores, Functional Analysis Through Hidden Markov Models (FATHMM) scores, probability of being loss-of-function intolerant (pLI), Combined Annotation Dependent Depletion (CADD) scores, and meta-voting prediction (MVP) scores between ET patients and controls for deletions and duplication separately using logistic regression. No significant associations between ET status and any of the CNV pathogenic prediction scores were identified (Figure 5).

**Figure 5.**
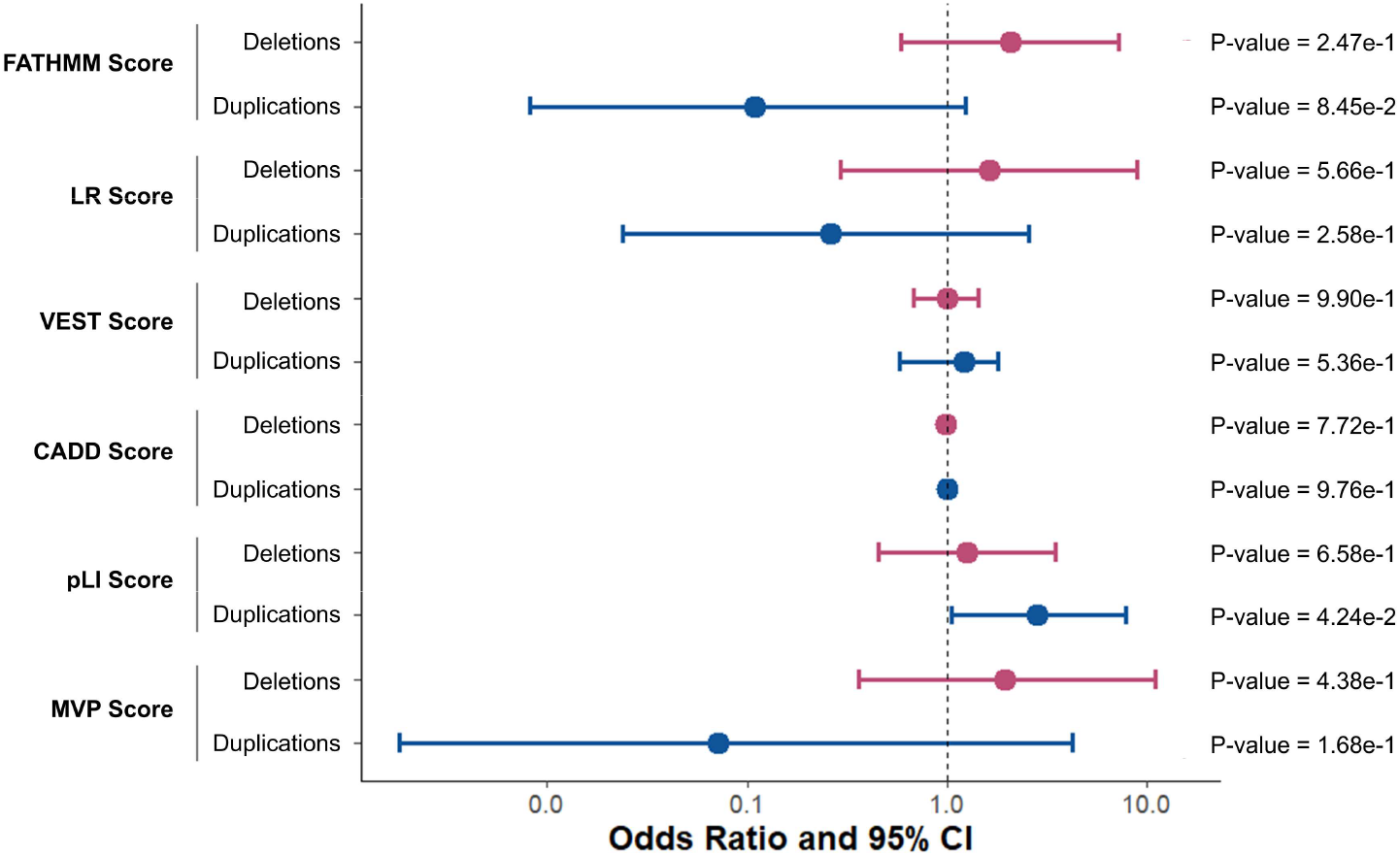
Pathogenic prediction scoring burden analyses of rare copy number variants encompassing protein coding regions. Forest plot depicting logistic regression results from tests of ET status with average pathogenicity score burden associations of Functional Analysis Through Hidden Markov Models (FATHMM), logistic regression (LR), Variant Effect Scoring Tool (VEST3), Combined Annotation Dependent Depletion (CADD), Probability of being Loss-of-function Intolerant (pLI), and meta-voting prediction (MVP) scores are shown. Odds ratios and 95% confidence intervals for each burden metric regression analyses are shown in red for deletions and in blue for duplications. P-values are also displayed. Bonferroni adjusted p-value cut-off for deletions and duplications: 0.05/12.

## Discussion

Despite being highly heritable, there is a disparity between the heritability of ET and explanatory genetic risk factors.^2,6^ With this in mind, and knowing that several studies have demonstrated that CNVs can explain the diagnoses of individuals with movement disorders, we posited that largely unexplored variant types, such as CNVs, might be a significant contributor of ET genetic risk.^9,10,13,40^ Here, we have conducted the largest study to date of CNVs in ET in a cohort of 1,853 ET patients and 10,336 unaffected controls. We identified rare CNVs encompassing protein coding regions, including 143 deletions and 180 duplications in cases, and 1,348 deletions and 1,709 duplications in controls. Though we could not definitively resolve any new candidate genes or replicate previous ones, our findings suggest that duplications may be of particular interest in ET, given their significantly higher prevalence in cases, greater overlap with Mendeliome and brain relevant genes, and greater burden in genomic disorder regions.

At the global level, ET cases showed a significantly greater burden in the number of duplications, the total duplication length, and the number of genes affected by duplications compared to controls. Further subsetting of CNVs by size revealed significant associations between the total number of duplications and the number of genes encompassed by duplications in the size range of 100-500 kbs with ET status. This suggests that duplications 100-500 kbs in size drove the global burden results seen across duplications. It is worth noting that duplications typically in this same 100-500 kbs size range have been reported in other later onset neurological disorders such as autosomal dominant leukodystrophy, making modest sized duplications a plausible disease mechanism for ET as well.^41^

At the global gene-level, burden tests revealed that the *ZNF813* gene was affected by significantly more duplications in controls than in cases, suggesting a potential protective factor for ET. The *ZNF813* gene is expressed in the brain and has been suggested to be associated with Coenzyme Q10 Deficiency Disease.^39,42^ Given its role in managing oxidative stress, Coenzyme Q10 is interesting in the context of ET since a recent linkage study suggested that genes involved in reactive oxygen species pathways may be relevant to ET.^43^ Interestingly, though the *ZNF813* gene is predicted to be a transcription factor,^42^ it sub-cellularly localizes with vesicles in human cell lines according to the Human Protein Atlas.^39^ When it comes to the cellular expression in the brain, the Human Protein Atlas also predicts that *ZNF813* clusters with oligodendrocytes from single cell data,^39^ which is a cell type proposed to be involved in the pathology of ET.^44^ Across data from a recent single-nucleus RNAseq study of the cerebellar cortex in ET patient, *ZNF813* was detected in microglia and oligodendrocyte clusters, albeit with low expression, which is consistent with what we expect given that our duplication results suggest a protective role of this gene in ET.^44^ Future *in-vivo* studies may be interesting to observe if a *ZNF813* duplication could rescue the ET phenotype in animal models. Additional *in vivo* or *in vitro* testing to elucidate the function of the *ZNF813* gene including monitoring its sub-cellular localization, measuring any reactive oxidative changes should coenzyme Q10 be involved, and observing its effects on oligodendrocyte function may be of interest to identify a potential role in ET. However, as Fisher’s exact tests were used in the gene-wise burden analyses to account for limited power at the gene-level, covariates could not be accounted for in these analyses. As such, the *ZNF813* finding should be interpreted with caution until otherwise functionally validated or replicated. These power limitations potentially also extend to the missed detection of differences between cases and controls elsewhere, given our focus on rare events.

The gene-set enrichment testing saw duplications affecting Mendeliome genes, genes highly expressed in the brain, and cerebellar genes as significantly associated with ET status. Interestingly, the *ZNF813* gene identified in the global gene-wise burden tests also belonged to the cerebellum expressed gene-set, though instead of being more prevalent in ET patients, it was more prevalent in controls and thus does not explain the positive association of duplications across genes expressed in the cerebellum with ET. These findings suggest that duplications relevant to ET patients affect genes involved in monogenic disorders and genes with brain-specific expression, including those in the cerebellum, a key region involved in the pathophysiology of ET.^2^ However, due to power limitations, it remains unclear whether the observed associations are driven by specific duplicated genes or by the cumulative effect of duplications across gene-sets. Further analyses on a genomic and functional level are necessary to elucidate the key contributors to these associations.

When looking at CNVs in genomic disorder regions, ET patients possessed a significantly greater frequency of duplications in these regions compared to those outside these regions. This finding corroborates that duplications are likely biologically relevant in ET since they coincide with regions that have a biological track record of being disease relevant. Biologically, these regions are often made vulnerable to nonallelic homologous recombination due to the presence of segmental duplications and low complexity repeats.^45,46^ These genomic disorder regions are well-established loci with strong CNV-disease associations, which continues to support the proposition that duplications may be disease relevant in ET.^45^

Finally, there were no associations observed in the pathogenic prediction burden tests, meaning that differences in the pathogenic character of CNVs could not be resolved between ET patients and controls. This lack of association may reflect limited power or perhaps instead that the X-CNV predictions are not sufficiently fine-tuned to detect pathogenic differences associated with modest-sized CNVs in a complex and later onset disorder like ET.

In conclusion, our results suggest that rare duplications affecting protein coding regions play a role in explaining the missing heritability of ET. These duplications tend to be modest in size (100-500 kbs) and capture regions highly expressed in the brain and specifically, the cerebellum. However, improved power and resolution is required to pinpoint potential putative genes and resolve pathogenic mechanisms. Additionally, as we work to address the missing heritability of ET, other lesser explored sources of variation, such as small structural variants, epigenetic modifications, or variations in non-coding regulatory regions may be the key to unlocking the genetic underpinnings of the disorder, offering new avenues for understanding and treatment of ET.

## Limitations

In this study, the use of SNP array genotyping technology for the detection of CNVs poses two key limitations. The first is that we cannot reliably detect CNVs less than 30 kbs in length, limiting our ability to assess potential contributions of smaller CNVs that could still be interrupting ET relevant genes.^23^ We also could not accurately assess the breakpoints of the identified CNVs.^47^ This is particularly relevant for duplications, as the additional copy of the genomic material inserted in the genome could disrupt the function of other genes. However, we do not have the resolution to detect such disruptions through array technology.^48^ Alternative technologies such as long-read sequencing could remedy both of these challenges.^7,49^ However, this becomes a sizeable investment, especially considering that additional power, and thus larger sample sizes, are required. Finally, our study only examined individuals of European ancestry. Future analyses encompassing more diverse cohorts of individuals may reveal population-specific roles of CNVs in ET.

## Supporting information

eTables

eMethods

## Data Availability

Patient data cannot be shared because of privacy restrictions. Control data were accessed through CARTaGENE and are available to other researchers upon request via their data access procedures (https://cartagene.qc.ca/)

## Code availability

The publicly available code for analyses are available and explained in the eMethods supplementary file.

## Ethics Statement

The McGill University Health Centre Research Ethics Board approved this work (Reference number: IRB00010120).

## Acknowledgements

Thank you to the members of the Rouleau laboratory for their support in this work and the patients who made this work possible. This research has been conducted using data and biosamples from CARTàGENE (https://cartagene.qc.ca/en).

## Author Contributions

(1) Analysis: A. Design, B. Execution, C. Review and Critique, D. Data management; (2) Manuscript Preparation: A. Writing first draft, B. Review and Critique.

MM: 1A, 1B, 2A

CL: 1A, 1C, 2B

AAD: 1A, 1C, 2B

JPR: 1A, 1C, 2B

VV: 2B

FA: 2B

CEC: 2B

DS: 1D

^†^ FH: 2B

*CVG: 2B

^†^ EGM: 2B

^†^ HAN: 2B

^†^ JAG-A: 2B

^†^ FJJ-J: 2B

^†^ PP: 2B

*AR: 2B

*AR: 2B

^†^ GD: 2B

^†^ GK: 2B

SLG: 1C, 2B

SMKF: 1C, 2B

*PAD: 1C, 2B

*GAR: 1C, 2B

* Canadian sample contribution (70% from Quebec; 30% Western Canada)

† European sample contribution (73% from Spain; 27% Germany)

