## supplemental Tables and Figures for "Copy Number Variant Duplications Associated with Essential Tremor"

### Supplemental Figures and Tables:

ET patients and control individuals underwent quality control at the sample level and CNV level. The number of individuals passing each quality control parameter is shown in Table S1.

| Quality Control Parameter | ET Samples | Control Samples |
|---|---|---|
| Raw Data | 1,853 | 10,336 |
| European | 1,572 | 9,860 |
| Unrelated | 1,507 | 9,630 |
| Raw CNVs | 1,485 | 9,629 |
| Log R Ratio Standard Deviation | 1,261 | 9,582 |
| B Allele Frequency Standard Deviation | 1,242 | 9,549 |
| Waviness Factor | 1,204 | 9,549 |
| Fragmented Call Correction | 1,204 | 9,549 |
| Size (CNVs > 30kbs) | 1,150 | 8,132 |
| CNV Number per Sample | 1,092 | 7,974 |
| Quality Score | 618 | 6,485 |
| Rare | 572 | 6,155 |
| Blacklist | 536 | 5,828 |
| Consensus | 505 | 5,563 |
| DeepCNV | 407 | 4,870 |
| Protein Coding | 244 | 2,402 |

**Table S1: The number of ET and control individuals remaining after each quality control step.** Rare CNVs were defined by Gnomad V4 structural variant European allele frequencies < 1%. Consensus CNVs were CNVs that were called by both PennCNV and QuantiSNP algorithms.

The exact number of deletions and duplications which remain after each quality control step for ET cases and controls is shown is Table S2:

| Quality Control Parameter | Essential Tremor Patients | | Controls | |
|---|---|---|---|---|
| | Deletions Count | Duplications Count | Deletions Count | Duplications Count |
| Raw CNVs | 106,118 | 7,992 | 140,818 | 42,104 |
| Log R Ratio Standard Deviation | 50,508 | 7,070 | 133,247 | 42,360 |
| B Allele Frequency Standard Deviation | 49,833 | 7,036 | 131,109 | 42,253 |
| Waviness Factor | 42,328 | 6,197 | 130,404 | 41,823 |
| Fragmented Call Correction | 41,860 | 6,112 | 127,377 | 39,842 |
| Size | 16,021 | 3,688 | 27,401 | 14,207 |
| CNV Number per Sample | 9,011 | 3,393 | 15,483 | 13,473 |
| Quality Score | 701 | 419 | 8,399 | 5,635 |
| Rare | 618 | 336 | 7,348 | 4,783 |
| Blacklist | 576 | 280 | 7,031 | 3,937 |
| Consensus | 563 | 230 | 6,794 | 3,247 |
| DeepCNV | 405 | 207 | 5,096 | 2,842 |
| Protein Coding | 143 | 180 | 1,348 | 1,709 |

**Table S2: The number of total deletions and duplications which remain after each quality control step for ET cases and controls.** Rare CNVs were defined by Gnomad V4 structural variant European allele frequencies < 1%. Consensus CNVs were CNVs that were called by both PennCNV and QuantiSNP algorithms.

All rare CNVs mapped to protein coding regions that were retained for analysis are shown in Figure S1 based on chromosome and position, colored by disease status and CNV type.

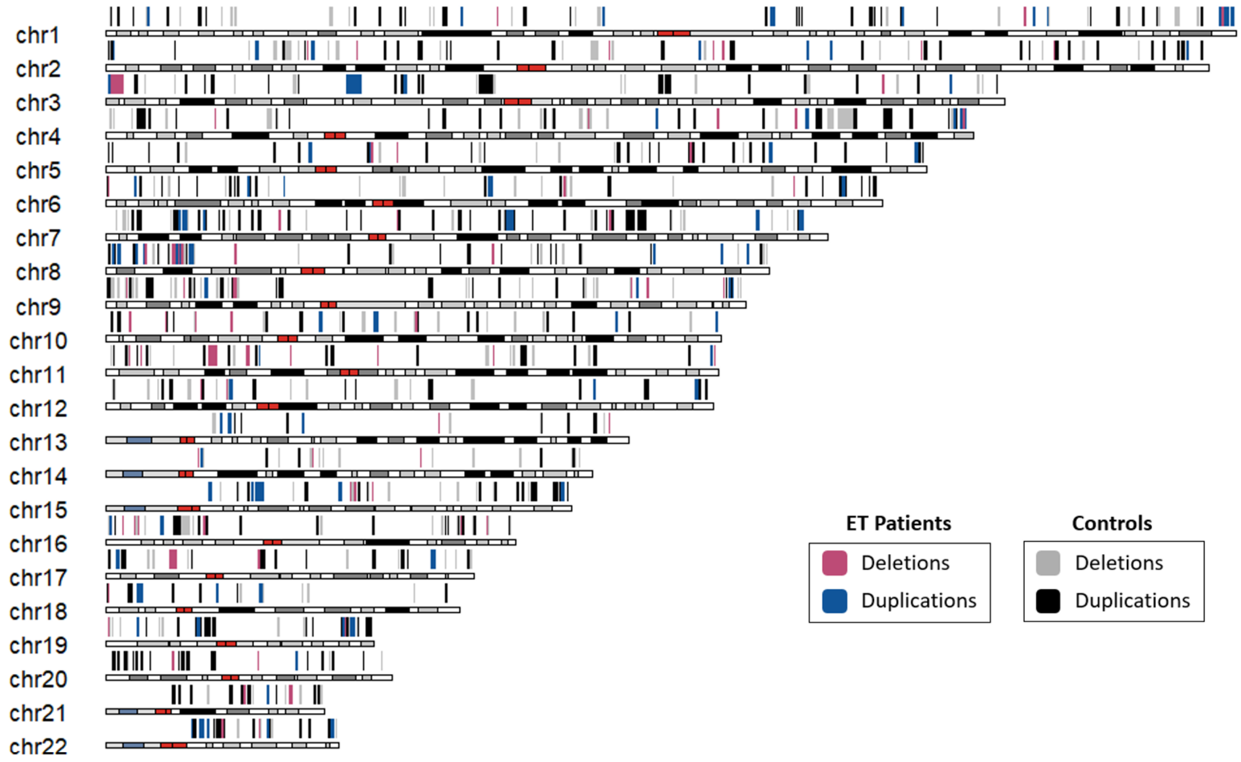


**Figure S1: Rare copy number variants mapping to protein coding regions shown across the genome.** Deletions and duplication are shown above the locus they map to per chromosome for ET patients and control individuals. ET patient deletions are shown by red boxes while ET duplications are shown by blue boxes. Deletions in the controls are shown by gray boxes while duplications in controls are shown by black boxes.

The ancestry of samples was inferred by principal component analysis (PCA) using 1000G as reference. The first two genetic principal components (PCs) are shown in Figure S2.

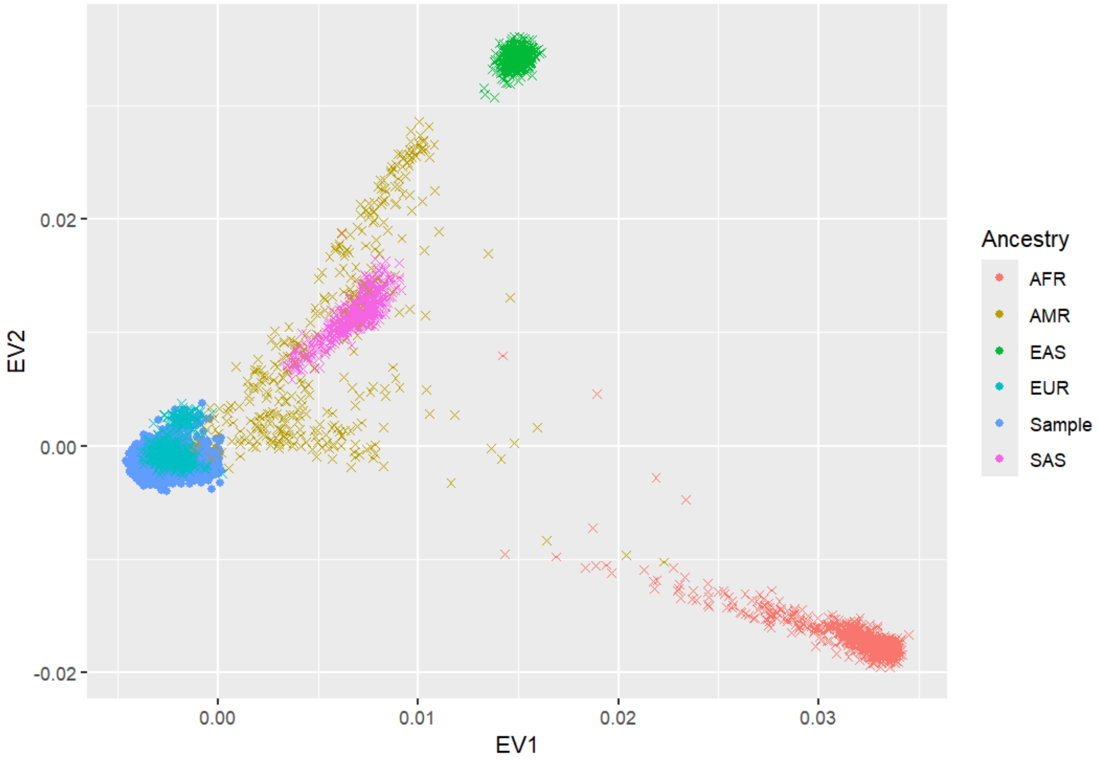


**Figure S2: Principal component analysis to infer sample ancestry.** Sample ancestry was estimated using principal component analysis (PCA) using PLINK v.1.9, with the 1000 Genomes Project as a reference panel. As 94% of samples clustered with European samples of the 1000 Genomes Project, only individuals with inferred European ancestry were included in the analysis. EV1 represents the first principal component while EV2 represents the second principal component.

The Scree plot capturing variance per principal component of the ancestry PCA is shown in Figure S3.

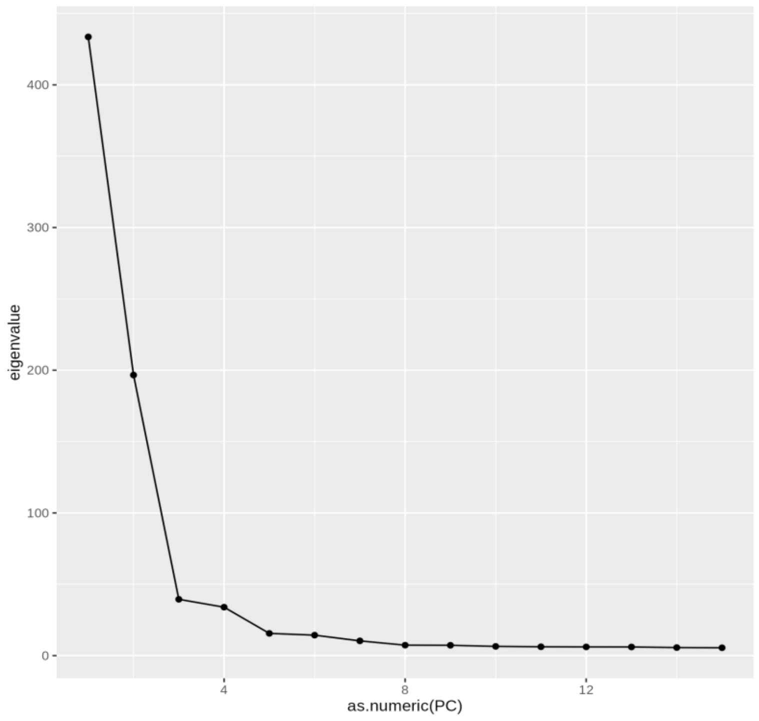


**Figure S3: Scree plot showing the variance explained by each principal component in the ancestry PCA analysis.** The x-axis represents the principal component number (PC) while the y-axis shows the eigenvalue which captures the variance explained by each PC.

A QQ-plot for gene-wise burden testing across deletions and duplication events was generated in R using the qqman() package shown in Figure S4. Notably, since we are testing the number of rare CNV events affecting a given gene, Fisher's exact test contingency tables are comprised of rather small counts which ultimately produce a limited set of possible p-values. This is reflected in the plateauing effect seen in Figure S4 as fewer tests (affected genes) and counts (number of CNVs) appear in rare event testing leading to limited, and thus several repeating, test statistic outcomes.

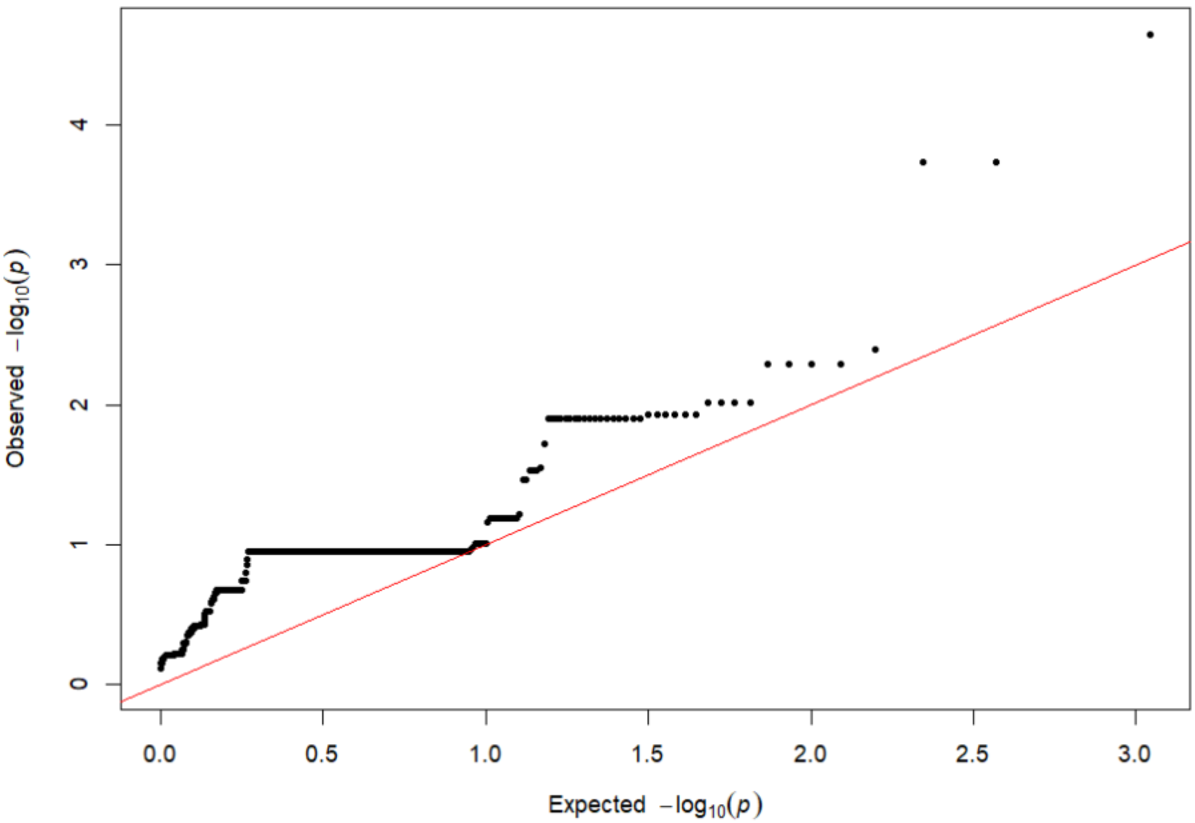


**Figure S4: QQ-plot of global gene-wise burden across deletions and duplications.** The diagonal red line illustrates the expected distribution under the null hypothesis. The plateauing of p-values reflects the limited resolution of tests with small CNV counts due to the focus on rare events.
