## Supplementary material for "Copy Number Variant Duplications Associated with Essential Tremor": eMethods

#### From the manuscript: *Copy Number Variant Duplications Associated with Essential Tremor*

### CNV Calling Pipeline

#### Step 1. Data Preparation

Begin by processing raw SNP array data in GenomeStudio as described previously. Export PLINK files for sample level quality control and for principal component calculation. Export Illumina report file for CNV calling

##### Sample level quality control (PLINK)

*Sample ancestry check:*

```
# Merge all cohort PLINK files and prepare for PCA:
## load module (PLINK2)
module load plink/2.00-20231024-avx2

## merge binary files with PLINK2:
#  plinks2merge.txt is a list of PLINK files with one file name per row
plink2 --pmerge-list plinks2merge.txt bfile --merge-max-allele-ct 2 \
--make-bed --out combined_cases_controls

# clean up the PLINK files:
plink --bfile combined_samples \
--memory 8100 \
--maf 0.05 \
--hwe 0.000001 midp \
--geno 0.02 \
--make-bed \
--out combined_snpQC

# Prepare PLINK files for PCA calculations
## Prune
plink --bfile combined_snpQC  --indep-pairwise 50 5 0.5 --out combined_snpQC_
prune

plink --bfile combined_snpQC  --exclude combined_snpQC_prune.prune.out \
--make-bed --out combined_snpQC_PC_prune
```

Test ancestry in R:

```
#USAGE: Rscript SNPRelate.PCA.R [prefix_of_plink_files]

#if first time, install as follows:
# install.packages("tidyverse")
# install.packages("BiocManager")
# BiocManager::install("SNPRelate")
# yes to update all

library(tidyverse)
library(SNPRelate)
```

```
#SAMPLES INPUTS
#import plink files
bed.fn <- "combined_snpQC_PC_prune.bed"
fam.fn <- "combined_snpQC_PC_prune.fam"
bim.fn <- "combined_snpQC_PC_prune.bim"

#combine plink inputs into GDS file
snpgdsBED2GDS(bed.fn, fam.fn, bim.fn, "sample.gds")

#merge the GDS files together
#1000G reference is used
snpgdsCombineGeno(gds.fn      = c("/ALL.chrALL.phase3.genotypes.maf05.LD.good
.gds", "sample.gds"),
                  out.fn      = "merged.gds",
                  same.strand = TRUE,
                  method      = "position")

#open the new merged gds file
merge.geno <- snpgdsOpen("merged.gds")

#compute PCA
#OPTIONS: minimum MAF, number of eigenvalues, number of threads
PCA <- snpgdsPCA(merge.geno,
                 maf         = 0.05,
                 eigen.cnt   = 15,
                 remove.monosnp = TRUE,
                 autosome.only = TRUE,
                 bayesian = TRUE,
                 num.thread = 8,
                 verbose     = TRUE,
                 missing.rate = 0.001)

#extract the sample ids and the top 15 PCs
table <- data.frame(sample.id        = PCA$sample.id,
                    EV1              = PCA$eigenvect[,1],
                    EV2              = PCA$eigenvect[,2],
                    EV3              = PCA$eigenvect[,3],
                    EV4              = PCA$eigenvect[,4],
                    EV5              = PCA$eigenvect[,5],
                    EV6              = PCA$eigenvect[,6],
                    EV7              = PCA$eigenvect[,7],
                    EV8              = PCA$eigenvect[,8],
                    EV9              = PCA$eigenvect[,9],
                    EV10             = PCA$eigenvect[,10],
                    EV11             = PCA$eigenvect[,11],
                    EV12             = PCA$eigenvect[,12],
                    EV13             = PCA$eigenvect[,13],
                    EV14             = PCA$eigenvect[,14],
```

```
                    EV15             = PCA$eigenvect[,15],
                    stringsAsFactors = FALSE)

#merge the 1kg PED with the dataframe
## supergroup file 1000G: integrated_call_samples_v2.20130502.ALL.supergroups
.ped
ped <- read.table("/integrated_call_samples_v2.20130502.ALL.supergroups.ped",
head=FALSE)
colnames(ped)[2] <- "sample.id"
colnames(ped)[6] <- "Ancestry"
table.ped.merge <- merge(x = table,
                         y = ped,
                         all.x = TRUE,
                         by = "sample.id")
# assign "Sample" to all non-reference entries
table.ped.merge$Ancestry[is.na(table.ped.merge$Ancestry)] <- "Sample"

#save the PCA table
write.table(table.ped.merge,
            file      = "PCA.SNPRelate.txt",
            quote     = FALSE,
            row.names = FALSE,
            sep       = "\t")

#generate eigenvalues and scree plot
scree <- as.data.frame(head(PCA$eigenval, n = 15))

scree <- rownames_to_column(scree)

colnames(scree) <- c("PC","eigenvalue")

write.table(scree,
            file      = "PCA.SNPRelate.scree.txt",
            quote     = FALSE,
            row.names = FALSE,
            sep       = "\t")

screeplot <- scree %>%
             ggplot(aes(x = as.numeric(PC), y = eigenvalue)) +
             geom_point() +
             geom_line()

ggsave(screeplot,
       filename = "PCA.SNPRelate.scree.tiff",
       device   = "tiff")

# plot the first 2 PC
# colour by ancestry with shapes different between reference and sample
plot <- ggplot(table.ped.merge, aes(x = EV1, y = EV2, color=Ancestry))
```

```
 + geom_point(size=1.5, shape=ifelse(table.ped.merge$Ancestry=="Sample",16,4))
)

#save the plot (can be any format)
ggsave(plot,
       filename = "PCA.SNPRelate.tiff",
       device   = "tiff")

#delete the created merged GDS (optional)
#unlink("merged.gds", force = TRUE)
#unlink("sample.gds", force = TRUE)
```

Inspect plots and remove samples which do not cluster with the majority of cohort (non-European samples removed in this case. Re-run the above script to confirm that you have isolated the samples of interest. Output should look like this:

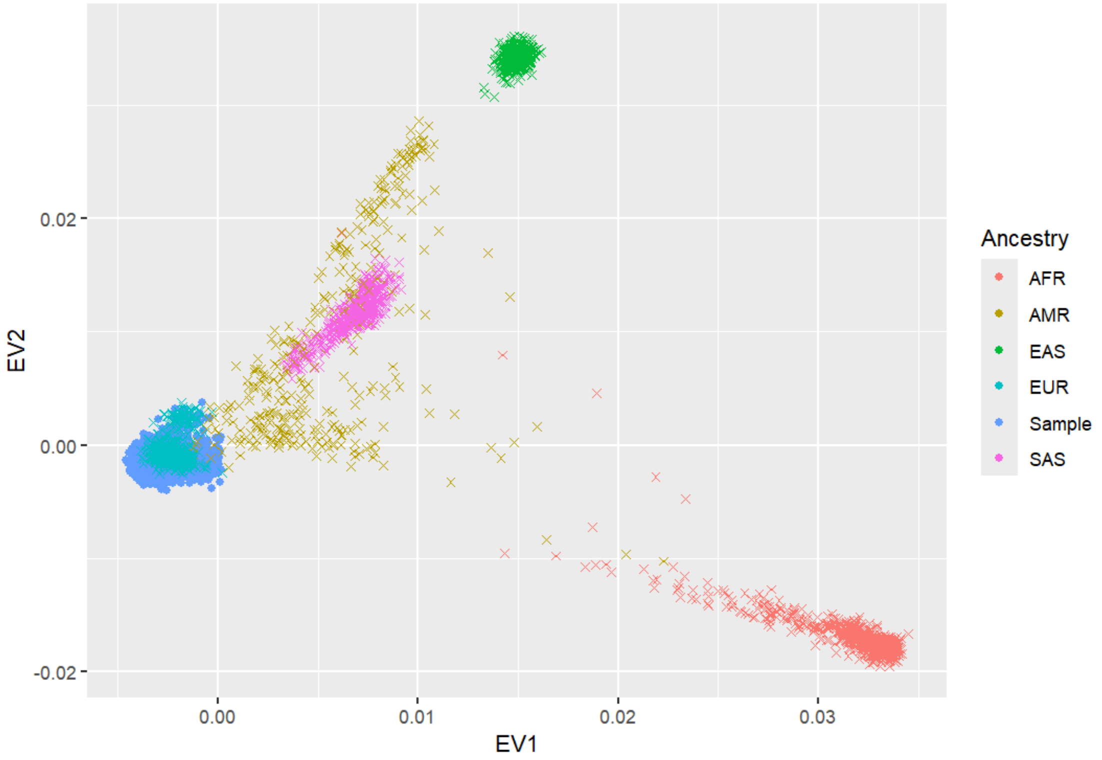

With accompanying screeplot:

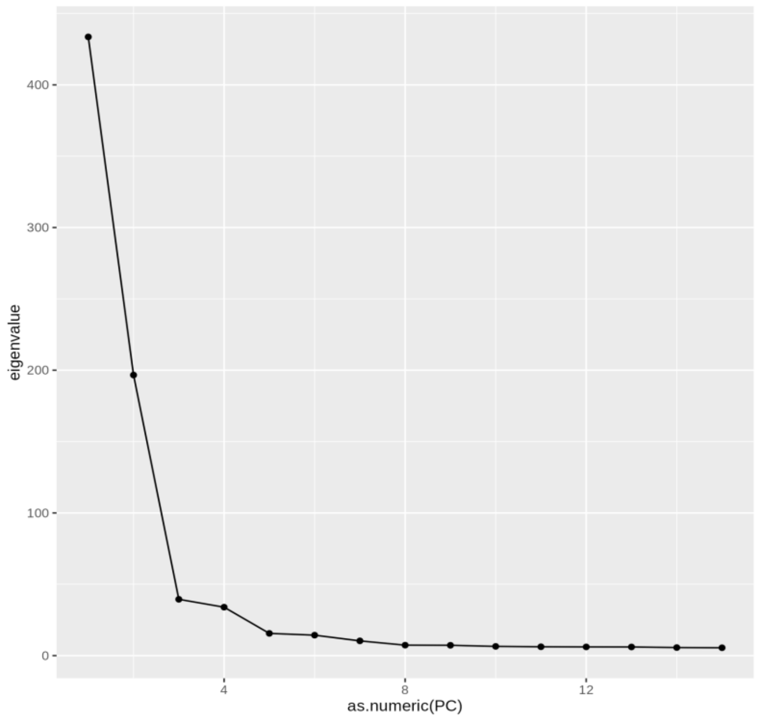

*Retain only list of unrelated individuals:*

```
# about output: --unrelated --degree 2 specifies that only unrelated pairs
# (up to the 2nd-degree in this case) between families are included
# in the output.
# more here: https://www.kingrelatedness.com/manual.shtml

king -b combined_snpQC.bed --kinship --unrelated --degree 2

# unrealted sample list in output: kingunrelated.txt
```

#### Step 2. PennCNV Calling

First install PennCNV: https://penncnv.openbioinformatics.org/en/latest/. To begin, signal intensity files must be generated from Illumina report files obtained in first step.

```
# prefix will be name ahead of all output files
# input_file is the Illumina Final Report file exported from GenomeStudio
./split_illumina_report.pl \
-prefix ./signal_intensity_files/${prefix}_ \
./starting_data/${input_file}
```

We must only consider SNPs in common between cohorts for CNV calling, so we must create a SNPfile with only those SNPs to use in PennCNV pipeline:

```
# from the individual cohort bim files, SNPs can be extracted
for file in *.bim; do awk '{print $2}' $file > ${file}_SNPs.list; done

# Run through the files to only retain SNP common to all files:
```

```
awk 'FNR==NR {count[$0]++; next} count[$0]==ARGIND-1 {count[$0]++} \
END {for (line in count) if (count[line]==ARGC-1) print line}' \
*_SNPs.list > CommonSNPs.txt
```

*Generate GC file in preparation for genomic wave adjustment:*

```
# this will generate the input for genomic_wave.pl --gcmodelfile

# module load penncnv/1.0.5
perl ./cal_gc_snp.pl <gc5Base_hg19.sorted.txt> <SNPfile> -output ET.GCmodel

# get GC file - be aware of your genome build:
wget http://hgdownload.cse.ucsc.edu/goldenPath/hg19/database/gc5Base.txt.gz
gunzip gc5Base.txt.gz
mv gc5Base.txt gc5Base_hg19.txt

# generate GC file:
perl ./cal_gc_snp.pl ./GC_content/gc5Base_hg19.txt CommonSNPs.txt -output ./c
ommon_ET.GCmodel
```

*Wave adjust with the newly generated GC file:*

```
# same $prefix as in signal intensity file generation - we are
# using all signal intensity files as input:
genomic_wave.pl ${prefix}_* --adjust --gcmodelfile ./common_ET.GCmodel \
--prefix ./wave_adjusted_signal_intensity_files/  --suffix adjusted
```

*Compute PFB file:*

```
# compile_pfb.pl will likely not accept all samples as input.
# Using just a few thousand is sufficient to accurately calculate PFB file.
# Make sure you are using only good quality samples in this computation.
# It is worthwhile to run this once with a blind selection of samples to be
# able to then extract sample level CNV QC parameters from detect_cnv.pl.
# Based on the naive QC summary of samples you obtain, you can select only
# those that pass QC cut-offs (more details in the CNV calling step) to
# feed into the compile_pfb.pl script

# randomly select 2000 samples from a set of good quality samples:
shuf -n 2000 list_good_samples.txt > good_samples_random2000.txt

# run pfb script:
compile_pfb.pl -list ./good_samples_random2000.txt \
-snpposfile ./CommonSNPs.txt -output case_control_2000.pfb
```

*Call CNVs:*

```
# make a file listing all wave-adjusted samples to call CNVs from wiht one fi
lename with path per line: list_WA_cases_controls.listfile
detect_cnv.pl -test -hmm $PATH/PennCNV-master/lib/hhall.hmm -pfb ./case_contr
ol_2000.pfb --listfile ./list_WA_cases_controls.listfile -conf -log test_wave
_adjusted.log -out test_wave_adjusted.rawcnv
```

Now we can commence additional sample level quality control based on CNV specific QC metrics:

```
# Extract QC metric from detect_cnv.pl log files:
# files will be in the format of: ID LRR_mean LRR_SD BAF_DRIFT WF
# input your own <path pointing to wave adjusted files> in grep command
grep "NOTICE: quality summary for <path pointing to wave adjusted files>" \
./test_wave_adjusted.log | \
sed 's,NOTICE: quality summary for <path pointing to wave adjusted files>,,g'
| \
awk '{print $1, $2, $4, $8, $9}' | sed 's/.adjusted//g' | sed 's/LRR_mean=//g
' | \
sed 's/LRR_SD=//g' | sed 's/BAF_DRIFT=//g' | sed 's/WF=//g' > \
test_ID_LRRmean_LRRsd_BAFdrift_WF.txt
```

Look at how the log R ratio SD, BAF drift, and WF parameters look. You'll want to visualize them to see the spread of these parameters to get a sense of the quality of the data. You'll want to remove outliers so you'll **calculate cut-offs of 3xSD for log R ratio SD and WF**. *BAF drift can be set to a static threshold of 0.002. If data looks really good and all samples would pass a 0.002 cut-off, a more strict 0.001 cut-off can be considered. Once these cut-off are calculated use filter_cnv.pl to clean up data. Keep in mind we are also filtering the data to only include CNVs that span **>= 10 SNPs (-numsnp)** and are **>= 30kbs in length (-length)**.

```
filter_cnv.pl ./test_wave_adjusted.rawcnv \
 -qclogfile ./test_wave_adjusted.log \
 -numsnp 10 -length 30k \
 -qclrrsd 0.2773094 \
 -qcbafdrift 0.002 \
 -qcwf 0.0347662 \
 -qcpassout ET_qcpass.list \
 -qcsumout ET_qcsum.list \
 -out ET_QC_LRRSD_BAF_WF_num_CNV
```

The ET_qcsum.list will leave you with a list of good quality samples from which samples for PFB file generation ought to be made from.

##### *Quality Scoring CNVs:*

This next step involves using the Mace et al. CNV QS algorithm. Source paper: A. Macé, M.A. Tuke, J.S. Beckmann, L. Lin, S. Jacquemont, M.N. Weedon, A. Reymond, Z. Kutalik, New quality measure for SNP array based CNV detection, *Bioinformatics*, Volume 32, Issue 21, November 2016, Pages 3298–3305, https://doi.org/10.1093/bioinformatics/btw477

This pipeline can be downloaded as follows in bash:

```
wget https://wp.unil.ch/sgg/files/2016/03/pennCNV_Pipeline_20160323.tar_.gz
```

To run the QS algorithm a config file must be generated specifying the location of PennCNV and your data. What actions you want the pipeline to take must be specified as well. Since CNV calling and all preceeding steps are already complete, we can enter later on into the pipeline. We will be using the pipeline to further clean the data and calculate QS for the CNVs. For CNV cleaning we will specifically be taking advantage of the *clean_cnv.pl combineseg* script included in the pipeline, which combines adjacent CNV calls which are predicted to be fragmented calls rather than two nearby independent CNVs. Once these adjacent CNV calls corrections are applied we will use the *filter_cnv.pl* script again, this time in the QS pipeline wrapper to filter out samples not just on our calculated LRR_SD, WF, and selected BAF_drift, but now that calls are corrected we can filter out samples which have too large of a number of CNVs i.e more than 150 CNVs (**-qcnumcnv 150**). This should be customized in the pipeline in the *filter_cnv.pl* line.

The config file that allows us to enter the QS pipeline looks like this:

```
pennCNVpath:    /home/medeiros/scratch/CLEAN_CNV_PIPELINE/tools/PennCNV-master
HMMpath:        /home/medeiros/scratch/CLEAN_CNV_PIPELINE/tools/PennCNV-master/lib/hh
all.hmm
HMMcreate:      0
PFB:    /home/medeiros/CLEAN_CNV_PIPELINE/PFB_generation/case_control_2000_QCd.pfb
CompilePFB:     0
GCmod: /home/medeiros/CLEAN_CNV_PIPELINE/GC_content/common_ET.GCmodel
UseGCmod:       0
InputData:      0
DATA:   /home/medeiros/CLEAN_CNV_PIPELINE/list_WA_cases_controls.listfile
OUTPUT: /home/medeiros/CLEAN_CNV_PIPELINE/PennCNV_calling_out/mace_QS_out
FormatedPath: /home/medeiros/CLEAN_CNV_PIPELINE/wave_adjusted_signal_intensity_files/sampleQCd/
Chromosome:     1-22
CNVcall:        0
Cleancall:      1
format: 0
CreateRfile:    1
AssoData:       0
NbCores:        1
PhenoPath:      /home/medeiros/CLEAN_CNV_PIPELINE/QS_mace/cases-controls_pheno.txt
Phenotype:      ET
```

The output directory specified by the config file (OUTPUT:) will contain the file with QS for each CNV. This can be easily filtered in R using the filter() command from the dplyr package. Good quality deletions should be selected as **QS < -0.5** and good quality duplications should be selected as **QS > 0.5**. CNVs satisfying these parameters will be extracted and saved to a file such as *CNVs_QS_0.5.txt* which will be used later for a starting point for filtering.

#### Step 3. QuantiSNP Calling

QuantiSNP is another CNV calling algorithm which will be used to identify consensus calls shared with PennCNV. Source paper: S. Colella, C. Yau, JM. Taylor, G. Mirza, H. Butler, P. Clouston, AS. Bassett, A. Seller, CC. Holmes, J. Ragoussis. QuantiSNP: an Objective Bayes

Hidden-Markov Model to detect and accurately map copy number variation using SNP genotyping data. Nucleic Acids Res. 2007;35(6):2013-25. doi: 10.1093/nar/gkm076.

QuantiSNP can be obtained as follows:

```
wget https://github.com/cwcyau/quantisnp/archive/refs/heads/master.zip
```

We will be using QuantiSNP through it's single file processing capability. Taken from the QuanitSNP "Howto":

> For single file processing, the input files must be plain tab-delimited text files with the following columns:

- Probe ID / SNP Name
- Chromsosome
- Position
- Log R Ratio
- B Allele Frequency

The already generated signal intensity files from PennCNV can be utilized for this.

```
# Provide a list of sample names to run in loop for single file processing:
list_of_samples=$1

# loop through list:
while read -r sample
do

# prepare sample file for input:
# make sure you are only including common SNPs in the analysis to be consiste
nt with the PennCNV analysis (use previously generated CommonSNPs.txt file to
retain only common SNPs)
awk 'BEGIN {FS=OFS="\t"} NR==FNR {a[$1]=$0; next} $1 in a {print a[$1], $0}'
\
/home/medeiros/CLEAN_CNV_PIPELINE/signal_intensity_files/*${sample}_* \
/home/medeiros/CLEAN_CNV_PIPELINE/commonSNPS/CommonSNPs.txt | \
awk '{print $1, $5, $6, $2, $3}' | sed 's/ /\t/g' | sed '1d' > ${sample}.txt

# correct header:
# header.txt should be one tab-delimited line that reads:
# Name    Chr    Position    Log R Ratio    B Allele Freq
cat header.txt ${sample}.txt > temp
mv temp ${sample}.txt

# get sample sex - this is a QuantiSNP input
# generate a text file (samples_sex.txt) with sample name in one column
# and sex in another (male/female) to be queried
sex=$(grep "$sample" ./samples_sex.txt | awk '{print $2}')
```

```
# run quantiSNP:
quantisnp --chr [1:22] \
--outdir /home/medeiros/scratch/CLEAN_CNV_PIPELINE/quantiSNP/output \
--sampleid $sample \
--gender $sex \
--emiters 10 --lsetting 200000 \
--gcdir /home/medeiros/scratch/CLEAN_CNV_PIPELINE/tools/QuantiSNP/b37 \
--plot --genotype \
--params params.dat --level levels.dat \
--input-files ${sample}.txt

# delete temp file:
rm ${sample}.txt

done < $list_of_samples
```

Each sample will generate an individual CNV file which can be concatenated with sample name to generate a list of CNVs called from QuantiSNP which can then be compared to the PennCNV calls.

#### Step 4. Filter CNVs

Now that CNVs have been called and the QS have been calculated we can begin filtering CNVs. Bedtools intersect (https://bedtools.readthedocs.io/en/latest/content/tools/intersect.html) will be used to identify which CNVs overlap with reference datasets.

To begin we filter the dataset to retain rare CNVs. This is done by extracting the common CNVs in GnomAD v4 from the non-euro SV database: https://gnomad.broadinstitute.org/data#v4-structural-variants

The rationale to use the common CNVs from GnomAD to filter rather than rare ones is that we are looking for CNVs in our own dataset which do not appear in the common subset of CNVs. If we looked at only rare CNVs from gnomAD and used that to intersect our dataset we might be inadvertently filtering out ultra rare CNVs not present in gnomAD. We thus identify the common deletions and duplications from gnomAD V4 as such, extracting for what is common in the European population to match our samples:

```
# get position, start, end, type, euro frequency, AF_nfe  columns
bcftools query -f '%CHROM %POS %END %SVTYPE %AF_nfe\n' \
gnomad.v4.1.sv.non_neuro_controls.sites.vcf > \
non_neuro_SV_chrom_pos_type_af.txt

## get with predicted introns - gene names
bcftools query -f '%CHROM %POS %END %SVTYPE %AF_nfe %PREDICTED_INTRONIC\n' \
```

```
gnomad.v4.1.sv.non_neuro_controls.sites.vcf > \
non_neuro_SV_chrom_pos_type_af_gene.txt

# select deletions:
grep "DEL" non_neuro_SV_chrom_pos_type_af.txt > non_neuro_DELs_chrom_pos_type
_af.txt

# select duplications:
grep "DUP" non_neuro_SV_chrom_pos_type_af.txt > non_neuro_DUPs_chrom_pos_type
_af.txt

# filter to get only common CNVs:
## in R:

library(dplyr)

df <- read.table("/home/medeiros/gnomadv4CNVs/reference/non_neuro_DELs_chrom_
pos_type_af.txt", header=FALSE)
colnames(df) <- c("CHR", "START", "END", "TYPE", "EUR_AF")

common <- filter(df, EUR_AF > 0.01)
write.table(common, file="euro_common_non-neuro_deletions_v4.txt", quote=F, r
ow.name=F, col.name=T, sep="\t")

# do same thing for duplications.

# these files are in build grch38, need to convert to hg19 for my dataset
```

Keep in mind that v4 of gnomAD is aligned to grch38 while my dataset is to hg19. So I had to lift over the file I just generated to hg19 using the UCSC server.

Filtering proceeds as follows using the dataset consiting of CNVs filtered by QS from earlier (CNVs_QS_0.5.txt) as the starting point. Bedtools takes BED files as input so data is transformed to be tab-delimited. Deletions and duplications are filtered separately here since they are separate events with seperate gnomAD references.

```
# begin filtering - we are filtering for CNVs that do not match the common CN
V reference:
# dels:
module load bedtools/2.29.2

bedtools intersect -a deletions_QS_0.5.BED -b common_deletions_gnomadv4_hg19.
bed \
-f 0.5 -r -v -wa > rare_deletions_QS_0.5.BED

bedtools intersect -a duplications_QS_0.5.BED -b common_duplications_gnomadv4
```

```
_hg19.bed \
-f 0.5 -r -v -wa > rare_duplications_QS_0.5.BED

#### now combine the two together to continue filtering:
cat rare_deletions_QS_0.5.BED rare_duplications_QS_0.5.BED > rare_CNVs_QS_0.5
.BED
```

Coding regions are then filtered (CNVs retained if they overlap any exon) - extracted from the UCSC Genome Browser:

```
bedtools intersect -a rare_CNVs_QS_0.5.BED \
-b ./protein_coding_RefGen_Hg19.BED -wa > protein_coding_rare_CNVs_QS_0.5.BED
```

Blacklist for hg19 is obtained from: https://github.com/Boyle-Lab/Blacklist/blob/master/lists/hg19-blacklist.v2.bed.gz. CNVs overlapping Blacklist are removed. Additionally hg19 centromeric regions defined by PennCNV as well as a 500,000 base buffer from chromosome starts and ends for telomeres were used:

```
# hg19 centromere positions (autosomal)
chr1:121500000-128900000
chr2:90500000-96800000
chr3:87900000-93900000
chr4:48200000-52700000
chr5:46100000-50700000
chr6:58700000-63300000
chr7:58000000-61700000
chr8:43100000-48100000
chr9:47300000-50700000
chr10:38000000-42300000
chr11:51600000-55700000
chr12:33300000-38200000
chr13:16300000-19500000
chr14:16100000-19100000
chr15:15800000-20700000
chr16:34600000-38600000
chr17:22200000-25800000
chr18:15400000-19000000
chr19:24400000-28600000
chr20:25600000-29400000
chr21:10900000-14300000
chr22:12200000-17900000

# hg19 telomere positions (autosomal)
# staring end:
chr1:1-500000
chr2:1-500000
chr3:1-500000
chr4:1-500000
chr5:1-500000
```

```
chr6:1-500000
chr7:1-500000
chr8:1-500000
chr9:1-500000
chr10:1-500000
chr11:1-500000
chr12:1-500000
chr13:1-500000
chr14:1-500000
chr15:1-500000
chr16:1-500000
chr17:1-500000
chr18:1-500000
chr19:1-500000
chr20:1-500000
chr21:1-500000
chr22:1-500000

# ending end:
chr1:248750621-249250621
chr2:242699373-243199373
chr3:197522430-198022430
chr4:190654276-191154276
chr5:180415260-180915260
chr6:170615067-171115067
chr7:158638663-159138663
chr8:145864022-146364022
chr9:140713431-141213431
chr10:135034747-135534747
chr11:134506516-135006516
chr12:133351895-133851895
chr13:114669878-115169878
chr14:106849540-107349540
chr15:102031392-102531392
chr16:89854753-90354753
chr17:80695210-81195210
chr18:77577248-78077248
chr19:58628983-59128983
chr20:62525520-63025520
chr21:47629895-48129895
chr22:50804566-51304566
```

Blacklist filtering bedtools command:

```
bedtools intersect -a protein_coding_rare_CNVs_QS_0.5.BED \
-b hg19-blacklist.v2.bed -wa | awk '!x[$0]++' > \
blacklist_protein_coding_rare_CNVs_QS_0.5.BED
```

Filter to consensus calls (retain CNVs in common with QuantiSNP):

```
bedtools intersect -a blacklist_protein_coding_rare_CNVs_QS_0.5.BED \
-b all_sample_CNVs_quantiSNP.txt -f 0.5 -r -wa | awk '!x[$0]++' > \
bothCallers_blacklist_protein_coding_rare_CNVs_QS_0.5.BED
```

#### Step 5. CNV Plotting + Deep CNV + Brief Manual Inspection

At the final step in the pipeline, remaining CNVs are plotted using PennCNVs *visualize_cnv.pl* script:

```
perl ./visualize_cnv.pl -format plot \
-signal /home/medeiros/CLEAN_CNV_PIPELINE/wave_adjusted_signal_intensity_file
s/sampleQCd/${Sample} \
/home/medeiros/CLEAN_CNV_PIPELINE/PennCNV_calling_out/mace_QS_out/all_QC_pass
ing_CNVs.rawcnv \
--snpposfile /home/medeiros/CLEAN_CNV_PIPELINE/commonSNPS/CommonSNPs.txt
```

A custom input file was generated for DeepCNV with the following columns (header):

```
echo "ID,label,Numsnp,Length,CN,Confidence,Number of CNVs in Sample,LRR_mean,
LRR_SD,BAF_mean,BAF_SD,BAF_DRIFT,WF,GCWF,CallRate,length_ind" > ET_all_CNV_sa
mples.csv
```

DeepCNV was then run as follows:

```
cd /home/medeiros/scratch/CLEAN_CNV_PIPELINE/deepCNV/DeepCNV-master

module load StdEnv/2020  gcc/9.3.0

module unload mugqic/python/3.9.1

module load python/3.8.10

module load opencv/4.5.5

export OMP_NUM_THREADS=4

export TF_NUM_INTRAOP_THREADS=4

export TF_NUM_INTEROP_THREADS=2

python  run.py /home/medeiros/scratch/CLEAN_CNV_PIPELINE/LRR_BAF_plots ET_all_C
NV_samples.csv ./output ./DeepCNV.hdf5
```

CNV output into the “good” directory were deemed high quality and were retained.

Manual inspection consisted of examining  LRR and BAF plots to ensure only high quality trustworthy CNVs were.

An example of a deletion that would be discarded for being of poor quality at this stage would have a LRR and BAF like this:

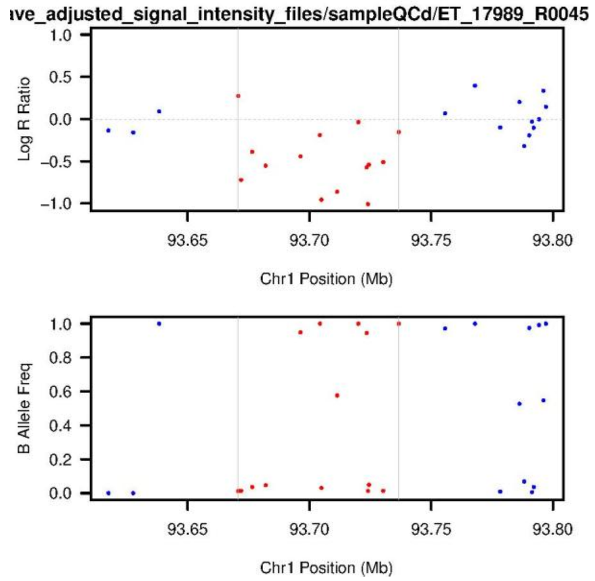

While a good quality deletion LRR and BAF would look like this:

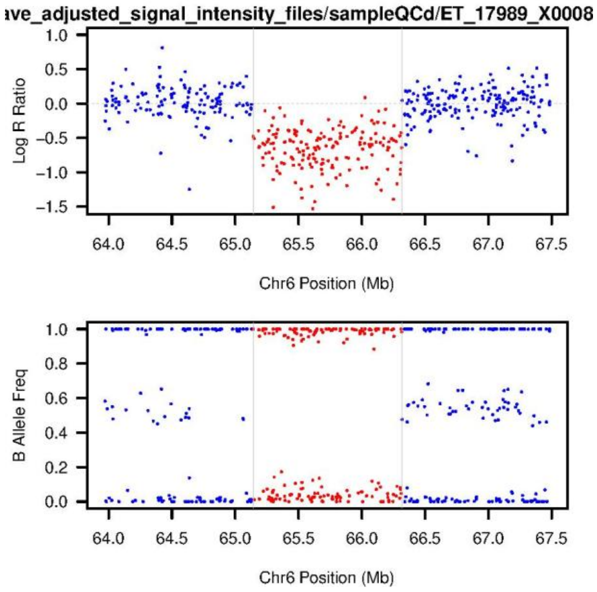

A bad quality duplication LRR and BAF plot may look like this:

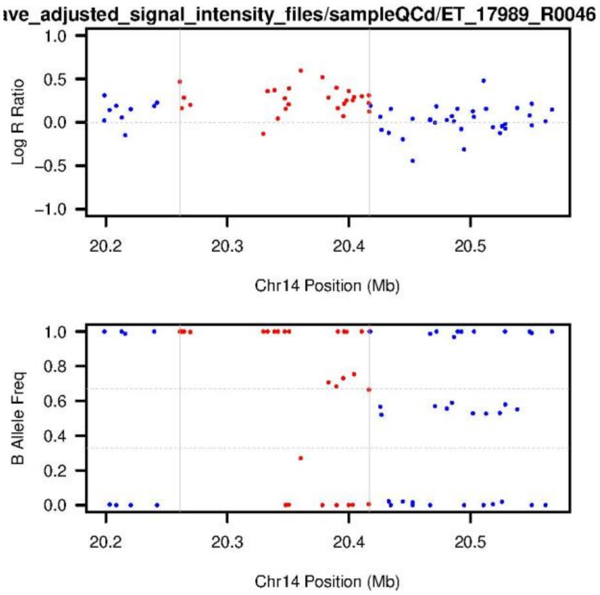

Whereas a good duplication LRR and BAF plot will look like this:

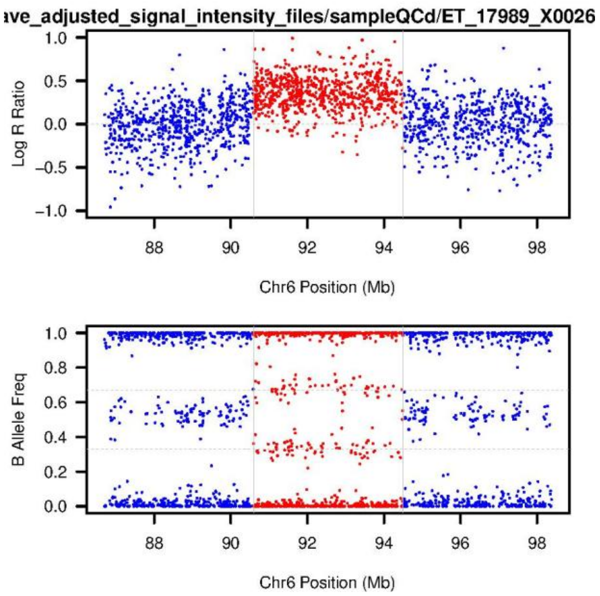

### Statistical Analysis

These analyses are done separately for deletions and duplications.

#### Global Burden

Global burden was tested across deletions and duplications separately for the following burden metrics:

- Number of CNVs

- Number of Genes affected by CNVs
- CNV length

These metrics were summed at the individual level to generate dataframe where each sample is a row and each column is an element to be used in the logistic regression. An example of this dataframe built for logistic regression is shown here (gene = gene affected by cnvs count; cnv = number of cnvs; size = total length of cnvs):

| ID | gene | cnv | size | LRR_SD | PC1 | PC2 | PC3 | PC4 | PC5 | PC6 | PC7 | PC8 | PC9 | PC10 | Status | sex |
|---|---|---|---|---|---|---|---|---|---|---|---|---|---|---|---|---|
| 78 | 1 | 1 | 238713 | 0.0957 | 0.0021 | -0.002 | 0.0089 | 0.0056 | 0.0113 | -0.021 | 0.004 | -0.011 | -0.001 | -0.011 | CONTROL | Female |
| 41 | 1 | 1 | 171150 | 0.0803 | 0.0052 | 0.0018 | 0.0099 | -0.009 | 0.0024 | 0.0008 | -0.006 | 0.0001 | 0.0139 | 0.0179 | CASE | Female |
| 81 | 1 | 1 | 390910 | 0.1373 | 0.0046 | 0.0010 | -0.0002 | -0.005 | 0.0055 | 0.0074 | 0.0062 | -0.06 | -0.002 | -0.004 | CONTROL | Male |
| 11 | 2 | 2 | 292657 | 0.1451 | -0.005 | -0.004 | -0.010 | 0.0168 | 0.0165 | -0.004 | -0.003 | 0.001 | 0.0024 | 0.0039 | CASE | Female |

Regressions were conducted in R as follows:

```
# run regression where burden_metric is either :
fit <- glm(as.factor(Status) ~ burden_metric + as.factor(sex)
           + PC1 + PC2 + PC3 + PC4 + PC5 + PC6 + PC7 + PC8 + PC9 + PC10
           + LRR_SD, data=all_together, family = binomial)

# extract OR, confidence intervals, and p-values:
exp(cbind(OR = coef(fit), confint(fit)))
coef(summary(fit))[,4]
```

The burden test by size segregation was done in the same way but data was subsetted based on size before burden metric counting as follows in R:

```
# load in data
df <- read.table("/home/medeiros/CLEAN_CNV_PIPELINE/final_CNVs_nice/CNVs_best
.txt" , header = TRUE)

# load library
library(dplyr)

# subset by size:
kb100 <- filter(df, Size < 100000)
kb500 <- filter(df, Size >= 100000 & Size < 500000)
MB1 <- filter(df, Size >= 500000 & Size < 1000000)
beyond <- filter(df, Size >= 1000000)
```

#### Pathogenic Prediction Burden

Pathogenic prediction burden is done for the most predictive coding and genome-wide pathogenicity scores from X-CNV: *Zhang, L., Shi, J., Ouyang, J. et al. X-CNV: genome-wide prediction of the pathogenicity of copy number variations. Genome Med 13, 132 (2021). https://doi.org/10.1186/s13073-021-00945-4*

LR scores, VEST3 scores, FATHMM scores, pLI, and CADD scores are annotated for each rare protein-coding CNV using X-CNV. X-CNV is installed as follows:

```
git clone https://github.com/kbvstmd/XCNV.git
cd XCNV
sh Install.sh
```

The input file is a tab delimited file describing rare protein coding CNVs through the following columns: Chr, Start, End, CNV Type.

Once annotation are made, the sum of predictive pathogenicity scores of all CNVs belonging to a given sample (∑Pathogenicity_metric) is calculated seperately for deletions and duplications for cases and controls. Example code of how counts are made for these metrics is provided:

```
# create a file listing all samples (one sample per row): list_of_samples.txt
# create a file which contains sample CNV data as well as X-CNV annotations.
# The format of this file should be one row per CNV-sample pairing along with
# accompanying X-CNV predictive scores. There should be one file for deletion
s
# and one for duplications. In this example we use: deletions_merged.txt

# build file of summed FATHMM, summed LR, summed VEST3, summed CADD, and summ
ed # pLI per sample. Each row of output (deleions_pred_sums.txt) will consist
of
# one unique sample:

# deletions:
while read -r sample
do
# query only lines matching to sample of interest:
grep "$sample" deletions_merged.txt > temp
# sum pathogenicity score metrics across sample individually:
FATHMM=$(awk -F' ' '{sum += $3} END {print sum}' temp)
LR=$(awk -F' ' '{sum += $4} END {print sum}' temp)
VEST=$(awk -F' ' '{sum += $5} END {print sum}' temp)
CADD=$(awk -F' ' '{sum += $6} END {print sum}' temp)
pLI=$(awk -F' ' '{sum += $7} END {print sum}' temp)
# append to file:
echo $sample $FATHMM $LR $VEST $CADD $pLI >> deletions_pred_sums.txt
done < list_of_samples.txt
```

Logistic regressions are then done in the same way as the global burden analysis but with ∑Pathogenicity_metric rather than burden_metric.

#### Gene-set Enrichment

Gene-set enrichment regressions are done much in the same way as the global burden analysis but instead the predictor is the number of genes belonging to a given gene-set which are affected by a sample's CNVs. The dataframe is again setup with one sample per row and each column being an element used in the regression. The same regression steps are taken in R. The Zinc Finger gene-set was assembled using biomaRT in R as follows:

```
# how to get Zinc Finger list in R:

# load library
library(biomaRt)

# Query all genes with descriptions:
genes <- getBM(attributes = c("hgnc_symbol", "description"),
               mart = ensembl)

# Filter genes with 'zinc finger' in their description
zinc_finger_genes <- genes[grepl("zinc finger", genes$description, ignore.cas
e = TRUE), ]
print(zinc_finger_genes)

# extract unique names - this is the gene-set list:
zinc_finger_list <- unique(zinc_finger_genes$hgnc_symbol)
```

#### Gene-based Burden

The gene-based regressions are done using a Fisher's exact test. To conduct the Fisher's exact test, first a dataframe must be built where one column contains the list of all genes that are mapped within the set of deletions or duplications from our cohorts, and the next column contains the number of instances a CNV affects that gene in case samples and in control samples. The dataframe should look like this example (there should be one table for deletions and one table for duplications):

| Gene | Number Cases with CNV Affecting Gene | Number of Controls with CNV Affecting Gene |
|---|---|---|
| ERCC8 | 1 | 0 |
| NBPF10 | 2 | 1 |
| MEI4 | 0 | 1 |

To do the Fisher's exact test, we need to fill in not just the number of samples that have a CNV affecting a given gene, but also we need to fill in the number of samples that *don't have* a CNV affecting a given gene. This can be done simply in R:

```
# Import dataframe:
counts_for_fishers <- read.table("~/final_CNVs_nice/counts_by_gene_dels.txt",
header=TRUE)

# modify column names:
colnames(counts_for_fishers) <- c("gene", "yes.cases", "yes.controls")
```

```
# Knowing how many samples have a CNV that affect a given gene and knowing
# the total number of samples that pass QC (ET cases: 1204 & Controls: 9549)
# we can fill in the number of cases and controls that do not have a CNV
# affecting a given gene as follows appending these counts to their own colum
ns:
counts_for_fishers <- mutate(counts_for_fishers, no.controls = 9549 - yes.con
trols)
counts_for_fishers <- mutate(counts_for_fishers, no.cases = 1204 - yes.cases)

# now order the columns so they're ready for input in our Fisher's function
# which will transform each row into a contigency table and run a Fisher's ex
act
# test for each row and save each gene's statistical output to a new row in a
# new output file. The order of the columns is important for the function
counts_for_fishers <- select(counts_for_fishers, "gene", "yes.cases", "yes.co
ntrols", "no.cases", "no.controls")

## DEFINE THE FISHER'S FUNCTION: ##########################

fishers_function <- function(x) {
    # extract the counts from row
    all_counts <- matrix(as.numeric(x[2:5]), nrow=2)

    # transform row into table using names
    rownames(all_counts) <- c("cases", "controls")
    colnames(all_counts) <- c("yes", "no")

    # do fishers exact test for the gene in that row
    output <- fisher.test(all_counts)

    # save all desired outputs to a list (order: gene, estimate, p.value, con
f.interval)
    final <- list(x[1], output$estimate, output$p.value, output$conf.int)

    # return output list
    return(final)
}

##########################################################

# now apply function to dataframe:

fishers_out <- apply(counts_for_fishers, 1, fishers_function)

# convert list into dataframe:
fishers_out_df <- as.data.frame(matrix(unlist(fishers_out), ncol=5, byrow=TRU
E))
```

```r
# add column names:
colnames(fishers_out_df) <- c("gene", "odds.ratio", "p.value", "conf.int_star
t", "conf.int_end")

# save:
write.table(fishers_out_df, "~/CNV_FINAL/deletions_fishers_exact_test.txt", q
uote = FALSE, row.names = FALSE, col.names = TRUE, sep ="\t")
```

Make sure to do this for deletions and for duplications separately because the events are not equivalent, they should be treated as distinct.

### Plotting

#### Plot Statistical results

Now that all CNVs have been called and statistics have been calculated for all tests we can visualize our statistics using a forest plot. This is an example of the code used to generate the forest plots in R:

```r
# load libraries:
library(ggplot2)
library(ggrepel)
library(ggpubr)
library(stringr)
library(RColorBrewer)

# set up dateframe with statistics filled in appropriately -
# keeping deletions and duplications as distinc entries:
results <- data.frame(
    tests = c("Number of Genes del", "Number of Genes dup",
                  "Number of CNVs del", "Number of CNVs dup",
                  "CNV Size del", "CNV Size dup"),
    OR = c(1.137545, 0.893,
           1.389111, 0.7280814,
            1.000001, 0.9999997),
    Lower_CI = c(0.9012739, 0.7221481,
                 0.5883705, 0.3534551,
                 0.9999993, 0.999999),
    Upper_CI = c(1.34567, 1.070321,
                 3.067538, 1.41091,
                 1.000002, 1.000001),
    p_val = c(1.83E-01, 2.62E-01,
                4.36E-01, 3.69E-01,
                2.02E-01, 5.44E-01)
)
```

```
# order cols of df - so they show up how we want in plot:
results$tests <- factor(results$tests,
                        levels = rev(c("Number of Genes del", "Number of Gene
s dup",
                                       "Number of CNVs del", "Number of CNVs
dup",
                                       "CNV Size del", "CNV Size dup")))

# plot:
ggplot(data = results, aes(x=tests, y=OR, ymin=Lower_CI, ymax=Upper_CI)) +
    geom_pointrange(aes(col = tests)) +
    geom_hline(yintercept = 1, linetype = 2) +
    xlab('Global Burden') + ylab('Odds ratio and 95% CI') +
    geom_errorbar(aes(ymin=Lower_CI, ymax=Upper_CI, col = tests), width=0.5,
cex=1) +
    coord_flip(clip = "off")+
    theme_classic() +
    theme(legend.position="none",
          legend.text=element_blank(), axis.title = element_text(size = 18, f
ace = "bold"),
          strip.text= element_text(size=16), axis.text.y = element_text(size=
10, colour = "black"),
          axis.text.x = element_text(size=16, colour = "black"),
          plot.margin = margin(1,6,1,1, "lines"), plot.title = element_text(s
ize=18)) +
    geom_text(aes(y=150,
                  label=ifelse(is.na(p_val), "",
                               paste0(" p = ", formatC(p_val, format = "e",
                                        digits = 2)))), size=5, hjust=0) +
    scale_y_continuous(trans="log10",labels = scales::number_format(accuracy
= 0.1)) +
    scale_x_discrete(labels = function(x) str_wrap(x, width = 25)) +
    scale_color_manual(values = list("#10559A", "#BD4C77", "#10559A", "#BD4C7
7", "#10559A","#BD4C77"))
```

This will generate a figure like this:

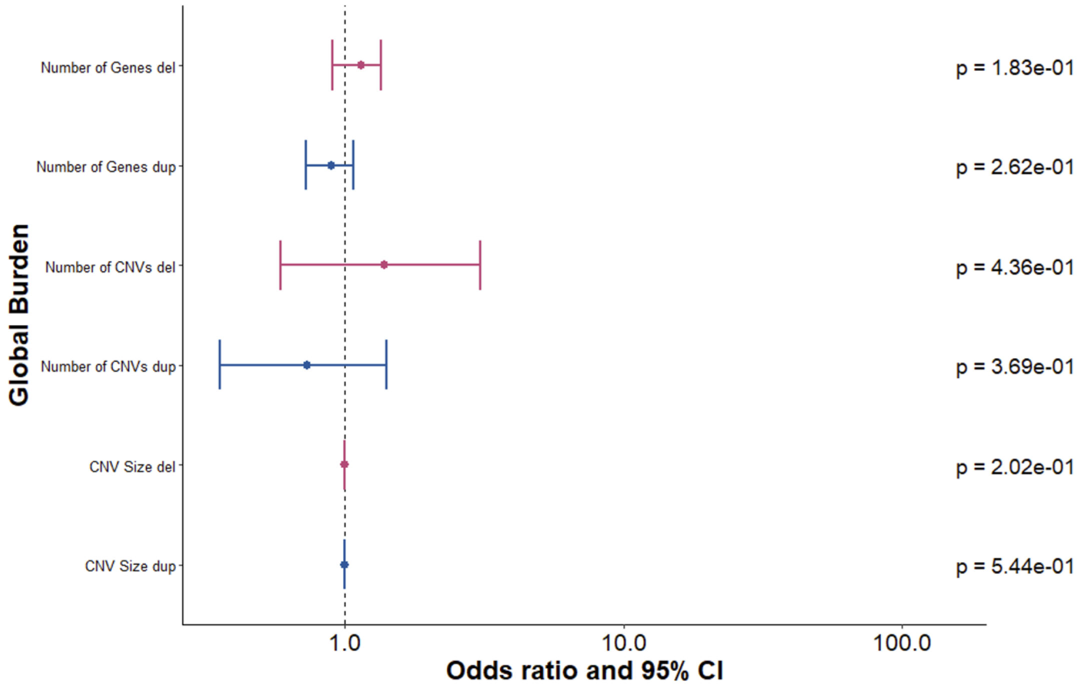

#### Plot CNVs

To see where all the CNVs happen to fall on the different chromosomes, we can create a dataframe listing all the CNVs with columns ‘chromosome’, ‘CN’, ‘start’, and ‘end’ for each CNV, where CN in the copy number and each CNV is a row. This visualization can be done in R:

```
# Load the necessary library
library(karyoploteR)
library(readr)

# read in your dataframe with columns
# 'chromosome', 'CN', 'start', and 'end' for each CNV.
# Here we assume that dataframe is called: CNV2plot

# setup base empty karyotype plot we will plot over
kp <- plotKaryotype(genome="hg19") # Use your reference genome

# loop through CNVs to plot
for (i in 1:nrow(CNV2plot)) {

# assign colors based on if deletion or duplication and also if case or contr
ol
color <- if (CNV2plot$CN[i] == "cn=3" & CNV2plot$Status[i] == "CONTROL") "bla
ck" else if
(CNV2plot$CN[i] == "cn=1" & CNV2plot$Status[i] == "CONTROL") "gray" else if
```

```
(CNV2plot$CN[i] == "cn=1" & CNV2plot$Status[i] == "ET") "#BD4C77" else if
(CNV2plot$CN[i] == "cn=3" & CNV2plot$Status[i] == "ET") "#10559A"

kpRect(kp,
       chr = CNV2plot$Chr[i],
       x0 = CNV2plot$Start[i],
       x1 = CNV2plot$End[i],
       y0 = 0.1, y1 = 0.9, # Set height of rectangles
       col = color, border = color)
}
```

This should generate a figure like this:

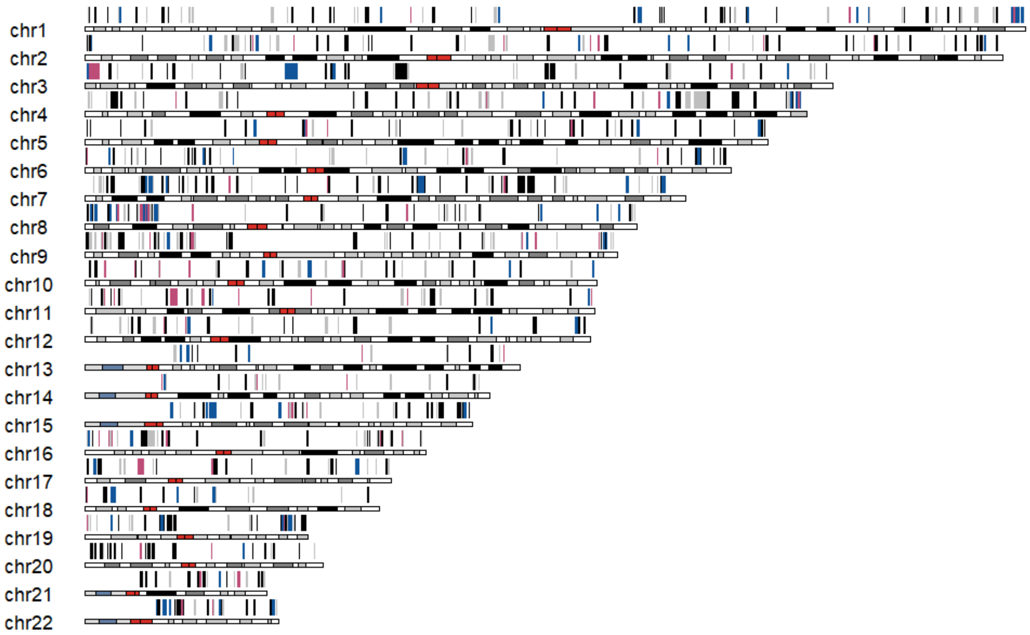

### Gene Sets

Description of gene-sets used in statistical analysis:

(1) Constrained genes from Gnomad v.2.1.1 (loss of function intolerant (pLI) genes: 3,036; constraint missense genes: 3,019).[1]

(2) Mendeliome genes: 3,204. We tested whether genes associated with mendelian inheritance showed enrichment in ET. The Mendel full gene set was downloaded from Breda Genetics (https://bredagenetics.com/) Sept. 2021 pulled (supplemental)

(3) Haplo-insufficient genes (ClinGen [Oct. 2022]: 329; rCNV: 2,987).

(A) ClinGen haploinsufficient data was pulled from the Clinical Genome Resource in October 2022.[2,3] Haploinsuficient genes identified from the from *Collins et al*.[4]

(B) Rare copy-number variants (rCNV) study were those with predict probabilities of dosage sensitivity (pHaplo) ≥ 0.86.

(4) Triplosensitive genes (ClinGen [Oct. 2022]: 15; rCNV: 1,559). ClinGen triplosensitive data was pulled from the Clinical Genome Resource in October 2022.[3] Triplosensitive genes identified from the from *Collins et al*.[4] rare copy-number variants (rCNV) study were those with predict probabilities of dosage sensitivity (pTriplo) ≥ 0.94.

(5) Neurodegenerative genes: 413. This list of genes was taken from the Adult onset neurodegenerative disorder (Version 4.38) panel (available from Genomics England: PanelApp https://panelapp.genomicsengland.co.uk/panels/474/). This panel is used for the clinical indication of 'R58 Adult onset neurodegenerative disorder' in the NHS Genomic Medicine Service and was originally created to merge following 7 panels: Hereditary ataxia (v1.148), Hereditary spastic paraplegia (v1.185), Early onset dystonia (v1.68), Parkinson Disease and Complex Parkinsonism (v1.64), Brain channelopathy (v1.46), Early onset dementia (encompassing fronto-temporal dementia and prion disease) (v1.45), and Amyotrophic lateral sclerosis/motor neuron disease (v1.26).

(6) Essential Tremor suggested genes: 37. The list of ET genes is as follows: *LINGO1, SLC1A2, STK32B, PPARGC1A, CTNNA3, FUS, HTRA2, TENM4, SORT1, SCN11A, NOTCH2NLC, NOS3, KCNS2, HAPLN4, USP46, CACNA1G, SLIT3, CCDC183, MMP10, GPR151, ETM1, DRD3, ETM2, ETM3, PRKG1, SAC3D1, SHF, TRAPPC11, NELL2, CACNA1A, PLCG2, ALDH3A2, CACNA1C, BACE2, LRRN2, DHRS13, LINC00323*. An overview of how these genes came to be proposed in ET is outlined *in Jimenez-Jimenez et al.*,[5] except for the *BACE2*, *LRRN2*, *DHRS13*, and *LINC00323* genes which were found to be associated with ET later in the *2022 Liao et al*.[6] paper which we also included in this gene set.

(7) The zinc finger genes list was extracted from the R package biomaRt[7,8] using Ensembl gene descriptions matching "zinc finger" as described earlier in eMethods.

(8) GISMO genes: 3,884. This list of genes was defined as the 1st and 2nd deciles of GISMO (Gene identity score of mammalian orthologs) constraint scores (lowest 20% of scores) taken from (https://www.biorxiv.org/content/10.1101/2024.05.16.594531v1). The GISMO scores assess constraint via gene loss across mammals weighed by evolutionary distance relative to humans.

(10) GISMO-mis genes: 3,136. This list of genes was defined as the 1st and 2nd deciles of GISMO-mis (GISMO-missense) constraint scores (lowest 20% of scores) taken from (https://www.biorxiv.org/content/10.1101/2024.05.16.594531v1). The GISMO-mis scores

assess constraint via the ratio of missense to synonymous variants across mammalian species for a given gene.

(11) sHet genes: 5,067. This list of genes was defined as the 8th to 10th deciles of sHet scores (top 30% of scores). The sHet score is a constraint measure derived from population genetics models and machine learning on gene features taken from (https://www.nature.com/articles/s41588-024-01820-9)

(12) Genes expressed in brain: 2,293. This set of genes was constructed using genes with either elevated expression in the brain, or exclusive expression in the brain as defined by the human protein atlas: https://www.proteinatlas.org/humanproteome/brain

(13) Synaptic genes: 1,602. This gene-set was constructed from the full set of genes defined in the SynGo (synaptic gene ontologies) 2023-12-01 release available here: https://syngoportal.org/

(14) Transcription factor genes: 1,637. This gene-set was constructed from the full set of genes in "TF list" download from the AnimalTFDB v4.0 dataset for homo sapiens, available here: https://guolab.wchscu.cn/AnimalTFDB4/#/Download
